# Morphometric Brain Networks at Ultra-Low Field: A First Assessment of Feasibility and Reliability

**DOI:** 10.64898/2026.10.17.26365791

**Authors:** Hajer Karoui, Niall J. Bourke, James H. Cole, Rui W. Yeow, Fred Dick, Raha Razin, David L. Thomas, Kirsten W.R. Schröder, David M. Cash, Martina Bocchetta, Steven C.R. Williams, František Váša

## Abstract

**Background:** Ultra-low field (ULF) magnetic resonance imaging (MRI) offers accessible point-of-care neuroimaging, yet its capability to support multivariate network analyses remains understudied.

**Methods:** We systematically evaluated the feasibility and reliability of Morphometric Similarity Networks (MSN) and Morphometric Inverse Divergence (MIND) networks derived from paired high-field (HF, 3T/1.5T) and ULF (64mT) scans in a primary cohort (N=23) and an independent replication cohort (N=19). We tested the impact of super-resolution processing, cortical parcellation granularity (Desikan-Killiany vs. Destrieux), and mesh resolution (fsnative vs. fsaverage5) on edge-level cross-scanner agreement (Lin’s concordance correlation coefficient, CCC) and subject-level identifiability.

**Results:** MIND networks demonstrated higher edge-level HF-ULF agreement (mean CCC = 0.95) and ULF reproducibility (mean CCC = 0.98) compared to MSNs (mean CCC = 0.86-0.87 and 0.91-0.94, respectively). In contrast, MSNs yielded higher subject-level differential identifiability across field strengths due to a broader distinction between within- and between-subject similarity. Reconstructing MIND networks on native high-resolution meshes (fsnative) maximised cross-scanner agreement, whereas coarser meshes and finer cortical parcellations enhanced individual network identifiability.

**Conclusions:** Structural similarity networks can be reliably constructed from ULF MRI data. Optimal analytic choices depend on study objectives: MIND on native meshes maximises cross-scanner stability, whereas MSNs excel at preserving individual uniqueness, paving the way for globally scalable connectomic research.

## 1 Introduction

Ultra-low field (ULF) magnetic resonance imaging (MRI) has emerged as a powerful technology for improving global access to neuroimaging. Its portability, low cost, and reduced energy requirements make it a suitable option for point-of-care applications and deployment in middle and low-resource settings, where high-field scanners remain unavailable, impractical, or scarce (Abate et al., 2024; Chetcuti et al., 2022; Vokhiwa et al., 2026). These systems have also been increasingly adopted in paediatric research, enabling studies of early brain development in communities that have been historically marginalised in neuroimaging research (Deoni et al., 2021; Cawley et al., 2023). Although the technology is relatively young, a growing body of work has demonstrated good correspondence with high-field (HF) MRI and high test-retest reliability across scanners and software versions (Váša et al., 2025; Hsu et al., 2025; Stockbridge et al., 2026; Sorby-Adams et al., 2024; Pretzsch et al., 2025; Cooper et al., 2024; Ringshaw et al., 2026). In parallel, advances in image reconstruction, image quality transfer, super-resolution, and segmentation tailored to ULF data have led to alternative pipelines for deriving quantitative morphometric measures at ULF (Krupinski et al., 2023; Deoni et al., 2022; Baljer et al., 2026; Baljer et al., 2025; Briski et al., 2026; Tapp et al., 2024; Iglesias et al., 2023; Billot et al., 2023b; Gopinath et al., 2026; Casella et al., 2026).

Taken together, existing work on ULF MRI provides a strong foundation for extending its use to complex multivariate analyses beyond regional volumetrics. Over the past decades, advances in neuroscience and MRI have introduced several approaches to mapping the brain as a large-scale network, collectively referred to as “connectomics”. Diffusion weighted imaging, for example, measures the directionally dependent water diffusion to reconstruct major white-matter tracts. Recent work by Gholam et al., 2026 demonstrated that such diffusion-based tractography at ULF is feasible in adults, but it currently requires long acquisition times, making it a less practical option especially when scanning vulnerable or young populations. Structural covariance network (SCN) analyses, on the other hand, use standard T1w MRI to derive networks based on morphometric similarity across regions and have been widely applied to study development, disease, and links to gene expression (Yao et al., 2010; Raznahan et al., 2011; Alexander-Bloch, Giedd, and Bullmore, 2013; Batalle et al., 2013; Kong et al., 2015; Li et al., 2017; Váša et al., 2018; Romero-Garcia et al., 2018; Seidlitz et al., 2018). While this approach is more practical than diffusion-based tractography, it was originally designed to model a single network per group of individuals, which limits the ability to capture idiosyncratic features and to study drivers of individual differences in brain networks. Although subject-level SCN methods have been proposed, approaches such as Morphometric Similarity Networks (MSNs; Seidlitz et al., 2018) and Morphometric Inverse Divergence (MIND) networks (Sebenius et al., 2023) were explicitly developed to construct subject-specific multivariate morphometric networks. These techniques build on histological and tract-tracing evidence linking coordinated cortical morphology to underlying axo-synaptic connectivity (Barbas, 2015; Goulas, Uylings, and Hilgetag, 2017; Goulas et al., 2016). MSNs are constructed from regional morphometric estimates and quantify the correlation of structural features between pairs of regions. In contrast, MIND networks use vertex-level morphometric estimates and compare the distributions of cortical features between regions via an inverse divergence measure, producing high-resolution similarity values that were shown to outperform MSNs (and diffusion-based tractography) in their correlation with a gene co-expression network and age-related changes (Sebenius et al., 2023; Sebenius et al., 2025). Recent work has shown that such morphometric similarity networks capture meaningful variation in cortical organisation across the lifespan and relate to cognition, gene co-expression, and neurodegenerative patterns (Tranfa et al., 2025; Yao et al., 2024; Wang et al., 2024; Fan et al., 2025; Han et al., 2026; Liang et al., 2025; Montagnese et al., 2025). Essentially, these approaches are built on a common theory, which states that regions sharing similar structural profiles are highly likely to be functionally connected. By being able to build a network map of physical structural similarities, we get a macroscale window into how the brain’s communication network is organised.

To date, however, MSN and MIND analyses have been restricted to HF MRI. With the growing adoption of ULF MRI, it remains unclear whether ULF data can support robust morphometric network reconstruction, and to what extent ULF-derived networks correspond to HF benchmarks. Despite their conceptual similarities, MSNs and MIND networks may behave differently when applied to ULF data. MSNs aggregate features at the regional level and might therefore be more robust to the lower spatial resolution and signal-to-noise ratio of ULF MRI, compared to the more fine-grained vertex-based MIND networks. At the same time, MIND offers a richer, higher-resolution characterisation of individual differences. Given the growing but HF-dominated literature on MSNs and MIND, it is not yet known which approach is better suited to ULF data, or whether widely used network construction pipelines can be directly translated to this setting. Addressing this question is important for guiding future methodological choices, and for leveraging ULF MRI in large-scale and globally distributed structural connectomic studies.

In this work, we systematically compare and evaluate the robustness of MSN and MIND networks derived from paired HF and ULF scans in healthy adults. Our aims were to: **(1)** quantify between-scanner agreement of MSN and MIND networks, **(2)** evaluate identifiability of inter-individual differences across field strengths and same-strength scanners, and **(3)** determine how parcellation choices (cortical atlas, mesh resolution) influence agreement and identifiability measures. To assess generalisability, we developed and optimised our analytic approach in a primary cohort, and then tested replication in an independent adult cohort scanned at a different site.

## 2 Methods

### 2.1 Participants

#### Primary cohort

23 healthy adults were recruited, aged 20–69 years, and balanced for age and sex (12 males and 11 females), with at least 2 males and 2 females per age decade. Exclusion criteria included major neurological and psychiatric conditions, conditions known to affect brain health, and any other contraindications to MRI. Participant screening was conducted using the standard MRI safety questionnaire and self-reported questionnaire regarding medical history. More information about this cohort can be found in Váša et al., 2025. The study was approved by the King’s College London Human Research Ethics Committee (Reference: HR/DP-21/22-26481).

#### Independent replication cohort

To assess the generalisability of our results, we analysed an independent cohort of 19 healthy adults aged 24–49 years (7 males and 12 females). Recruitment was an opportunity sample from within UCL. Exclusion criteria included major neurological and psychiatric conditions, conditions known to affect brain health, and any other contraindications to MRI. Participant screening was conducted using the standard MRI safety questionnaire and self-reported questionnaire regarding medical history. The study (“Comparative study of MRI quality at multiple field strengths”) was approved by the UCL PaLS LREC – EP – 2025-0815-608, F Dick PI.

### 2.2 MRI Data Acquisition

#### Primary cohort

Participants underwent 2 imaging sessions within a maximum of 36 days. The first session was performed at King’s College London’s Centre for Neuroimaging Sciences (CNS) on a 3T GE SIGNA Premier system, acquiring T1w MPRAGE and T2w sequences (1 mm³ isotropic). Immediately after, participants were scanned on a 64 mT Hyperfine Swoop® system using custom isotropic T1w and T2w protocols (T1w: 2.3 × 2.3 × 2.3 mm³; T2w: 2.2 × 2.2 × 2.2 mm³). Additionally, T1w and T2w scans were acquired using non-isotropic product sequences (T1w: 1.6 × 1.6 × 5 mm³; T2w: 1.5 × 1.5 × 5 mm³), each with higher in-plane resolution along the axial, sagittal, or coronal plane. The second session was undertaken at Evelina Newborn Imaging Centre (ENIC) on an identical Hyperfine Swoop® system with the same acquisition protocol and software version (rc8.6.0). All scans were acquired on both Hyperfine scanners, except for the isotropic T2w which was not acquired on the second session at ENIC. Scanning sessions were labelled as HF (3T), ULF_S1_ (Hyperfine at CNS), and ULF_S2_ (Hyperfine at ENIC). Pearltec inflatable pads were used in all three scanners to minimise head motion (Pearl Technology AG, Switzerland). For further details regarding the sequences, including acquisition times, see Váša et al., 2025.

#### Replication cohort

Participants were scanned on a clinical Siemens Magnetom Avanto 1.5T (VB17), and Magnetom Prisma 3T (VE11C), and 64mT Hyperfine Swoop® scanner (8.8.1). Data from all three scanners were acquired on the same day in counterbalanced order across participants. On the HF systems (1.5T and 3T), T1w and T2w images with isotropic resolution of 1mm³ were acquired. MR-MinMo (Mangal et al., 2026) head pads were used in both two scanners to minimise head motion. On the ULF scanner, acquisitions included a T1w sequence with 1.6 x 5 x 1.6 mm³ resolution, and three anisotropic T2w acquisitions with higher in-plane resolution along each orthogonal plane (T2w AXI: 1.5 x 1.5 x 5mm³; T2w COR: 1.5 x 5 x 1.5 mm³; T2w SAG: 5 x 1.5 x 1.5 mm³). Pearltec inflatable pads were used in the Hyperfine scanner. All scans passed quality checks and were included in the analyses. One subject was missing the T2w sequence on the 3T scanner. Scanning sessions were labelled as HF_3T_, HF_1.5T_, and ULF.

### 2.3 Preprocessing

In the primary cohort, each participant had 1 HF and 2 ULF scanning sessions. Since HF images were acquired at 1 mm³, no further pre-processing was applied before segmentation. For ULF scans, we used two image pre-processing pipelines to super-resolve the images in order to ensure they are isotropic and of comparable resolution and/or quality to the HF scans. We then tested the effects of each method on HF agreement and ULF reproducibility in the resulting brain networks. A diagram of the full processing pipeline can be found in Figure1.

In the first analysis branch, the three orthogonal non-isotropic T2w 64mT scans (axial, sagittal, and coronal) from the first ULF scanning sessions were combined into a single higher-resolution scan using multi-resolution registration (MRR) (Deoni et al., 2022). The resulting T2w MRR scan had an effective isotropic spatial resolution of 1.5 mm³, which was then resampled to 1 mm³. In the second analysis branch, we pre-processed the T1w isotropic scans (from each ULF session) with Supersynth (Iglesias et al., 2023; Billot et al., 2023a), to obtain synthetic T2w 1 mm³ isotropic scans. As previously mentioned, the T2w isotropic was not acquired in the second ULF session; we therefore used the T1w scan as input for super-resolution to ensure consistency across subjects and pipelines. Previous work by Váša et al., 2025 showed that, at 64mT, regional volume estimates derived from T2w images exhibit higher agreement with 3T T2w volumes than those derived from T1w images. We therefore used T2w images whenever an actual T2w acquisition was available (ULF_S1_), and relied on T1w scans only as input for generating synthetic T2w images for the second ULF session. Each of the T2w images (HF, ULF-MRR, and ULF-Synth) were then processed using a version-controlled Docker container of the FreeSurfer (version 8.1.0) recon-any tool for segmentation and cortical parcellation (Gopinath et al., 2026).

The replication dataset was pre-processed using the same pipeline as the primary cohort. Out of the 19 participants, one participant was missing a T2w scan on the 1.5T HF system. For this participant, analyses therefore included one HF and one ULF scan, whereas for all other participants, analyses included two HF (3T and 1.5T) and one ULF scan.

### 2.4 Network Construction

In this work, we compare two kinds of networks constructed using ULF scans: Morphometric Similarity Networks (MSN) and Morphometric Inverse Divergence (MIND) networks. As illustrated in Figure 2, Both are analysed as weighted networks, where the weight of an edge (connection) between two regions reflects the extent to which their morphometric features co-vary within a subject. One of the main differences between these two approaches is the spatial scale of the morphometric data used for network construction. MSNs are constructed from region-level average morphometric estimates (e.g., cortical thickness, surface area, volume etc.) within each parcel. For each subject, MSNs quantify the correlation across these structural features between pairs of regions. In contrast, MIND networks are constructed from vertex-level morphometric estimates, providing higher spatial resolution than the region-averaged features used in MSNs. For each subject, MIND edge weights reflect the similarity (inverse divergence) between the multivariate distributions of structural features across vertices between a pair of regions.

**Figure 1.**
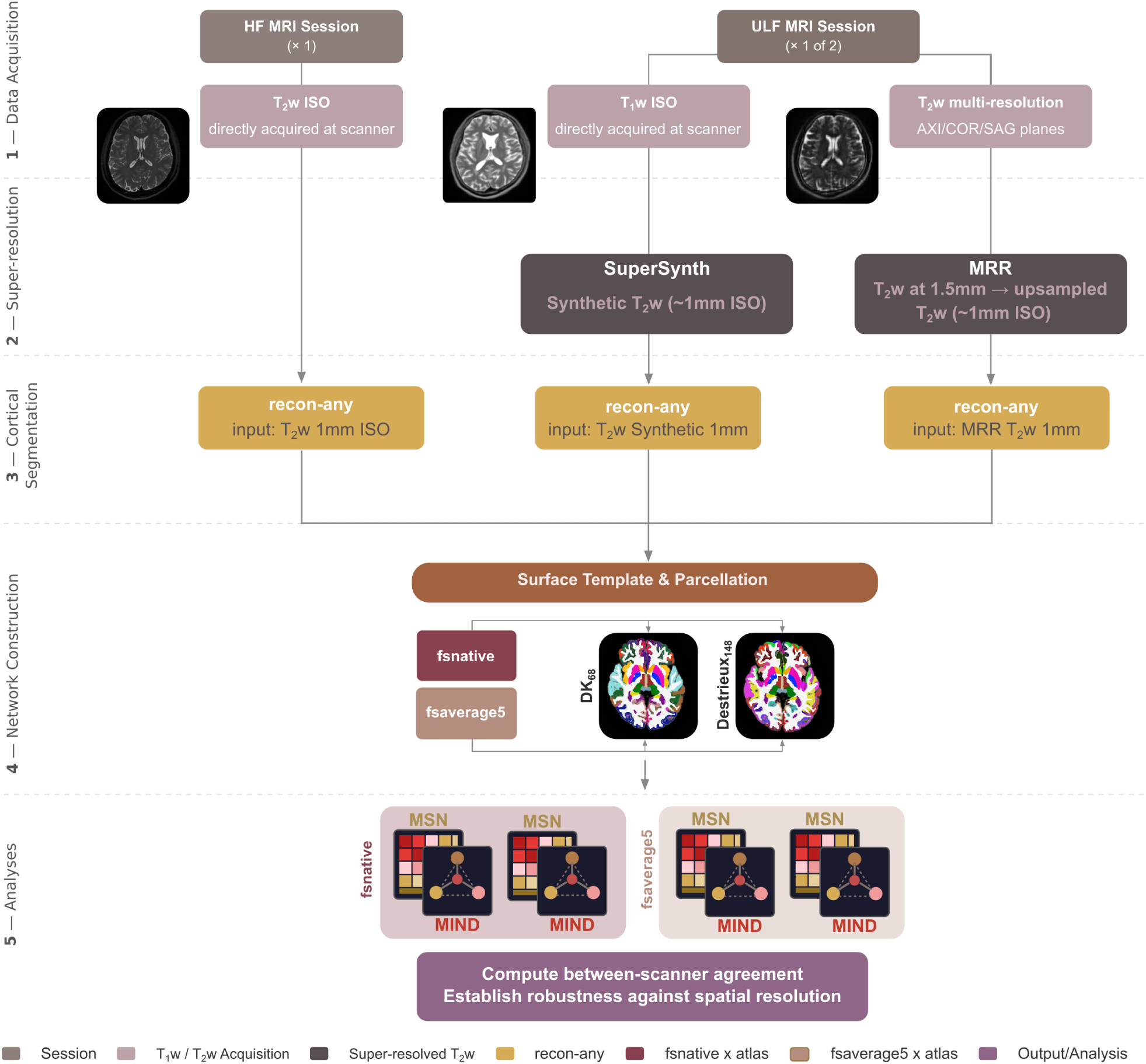
Analysis flow for super-resolution, cortical parcellation and segmentation, and network reconstruction per subject for the primary cohort.

**Figure 2.**
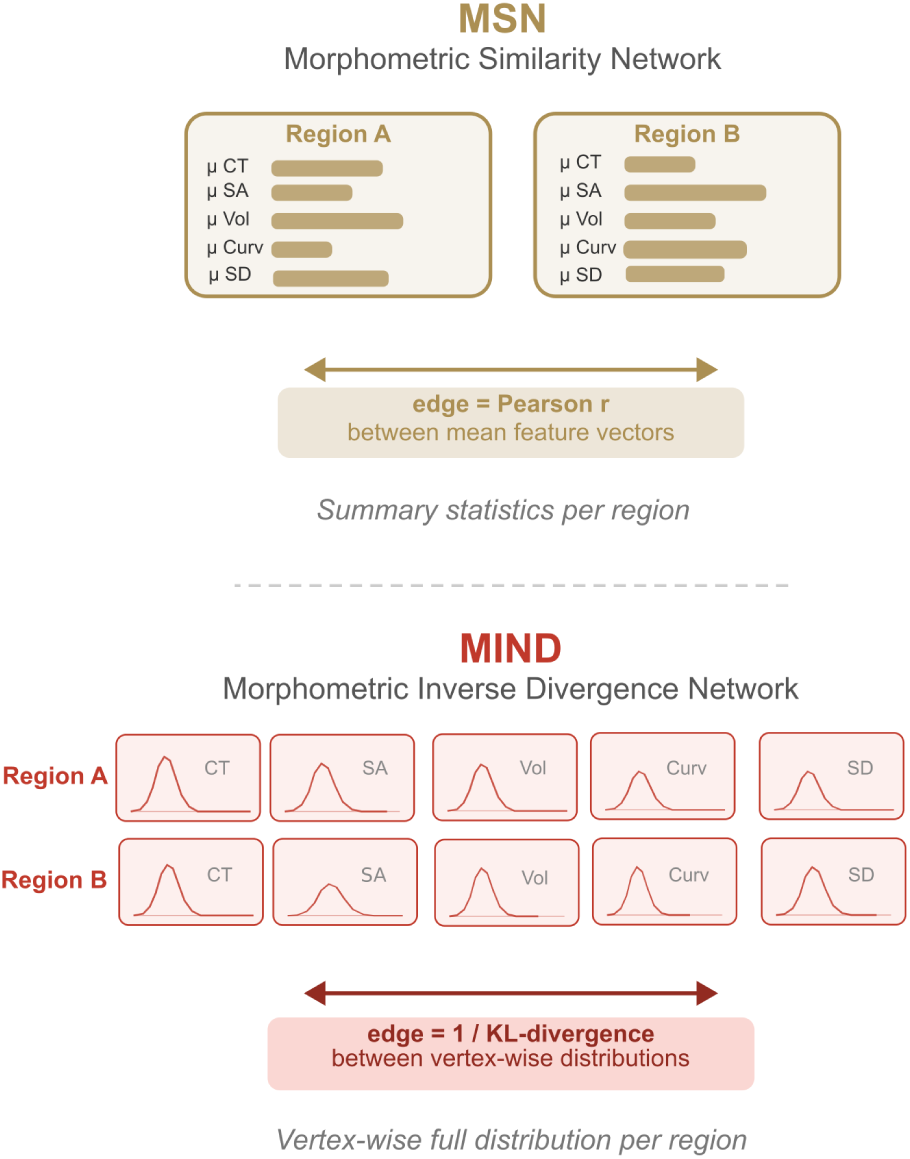
Network construction approaches comparing Morphometric Similarity Networks (MSN) and Morphometric Inverse Divergence (MIND) networks.

#### 2.4.1 MSN (Morphometric Similarity Networks)

For each subject and each scanning session, we built the *region* × *feature* matrix **X**, where for each region we have *N _f_* morphometric features (thickness, area, mean curvature, etc.):

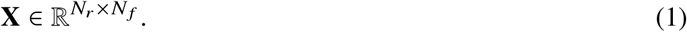

Row *i* represents the multivariate profile of region *i*, making it the **region feature vector**:

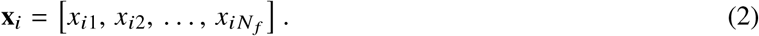

Column *j* represents the **feature map** for feature *j* over regions:

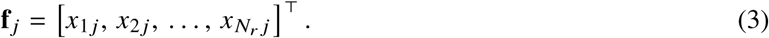

We then *z*-scored each **feature column** across regions and correlated the rows of the *z*-scored matrix pairwise:

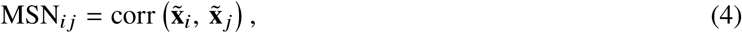

where **x̃***_i_* is the *z*-scored feature vector of region *i*.

The edges of a subject’s MSN are pairwise inter-regional Pearson correlations of morphometric feature vectors (Seidlitz et al., 2018). We applied this process using the Desikan-Killiany (N_ROIS_ = 68) and the Destrieux (N_ROIS_ = 148) cortical parcellations, for both the primary and replication cohorts.

#### 2.4.2 MIND (Morphometric Inverse Divergence Network)

MIND networks were constructed for every subject using the same five morphometric features as MSNs (Figure 3). As described in Sebenius et al., 2023, morphometric features were first z-scored to standardise values across the cortical surface. Vertices with values equal to zero were discarded to ensure only biologically plausible measures are used. The (dis)similarity between two regions with distributions *P*(*x*) and *Q*(*x*) was quantified using the Jeffreys-symmetrised Kullback-Liebler Divergence (KL) (Jeffreys, 1973). MIND networks were constructed using the open-source code made available by the authors, with minor modifications to allow changes in mesh resolution. As this method relies on vertex-level estimates of morphometric features, we applied the process described above using two mesh resolutions: *fsnative*, a high-resolution template, with the number of vertices depending on individual anatomy (up to ∼164,000 vertices), and *fsaverage5* which is a medium-resolution template containing 10,242 vertices per hemisphere for all subjects alike. This was achieved by resampling the subject’s fine mesh to the coarser *fsaverage5* mesh using FreeSurfer’s mri_surf2surf function, where the value of each coarse vertex is an interpolation of nearby fine-resolution vertices. We applied this process using the Desikan-Killiany and the Destrieux parcellations, for both the primary and the replication cohort. Structural features considered in network reconstruction include gray matter volume, sulcal depth, surface area, mean curvature, and cortical thickness.

**Figure 3.**
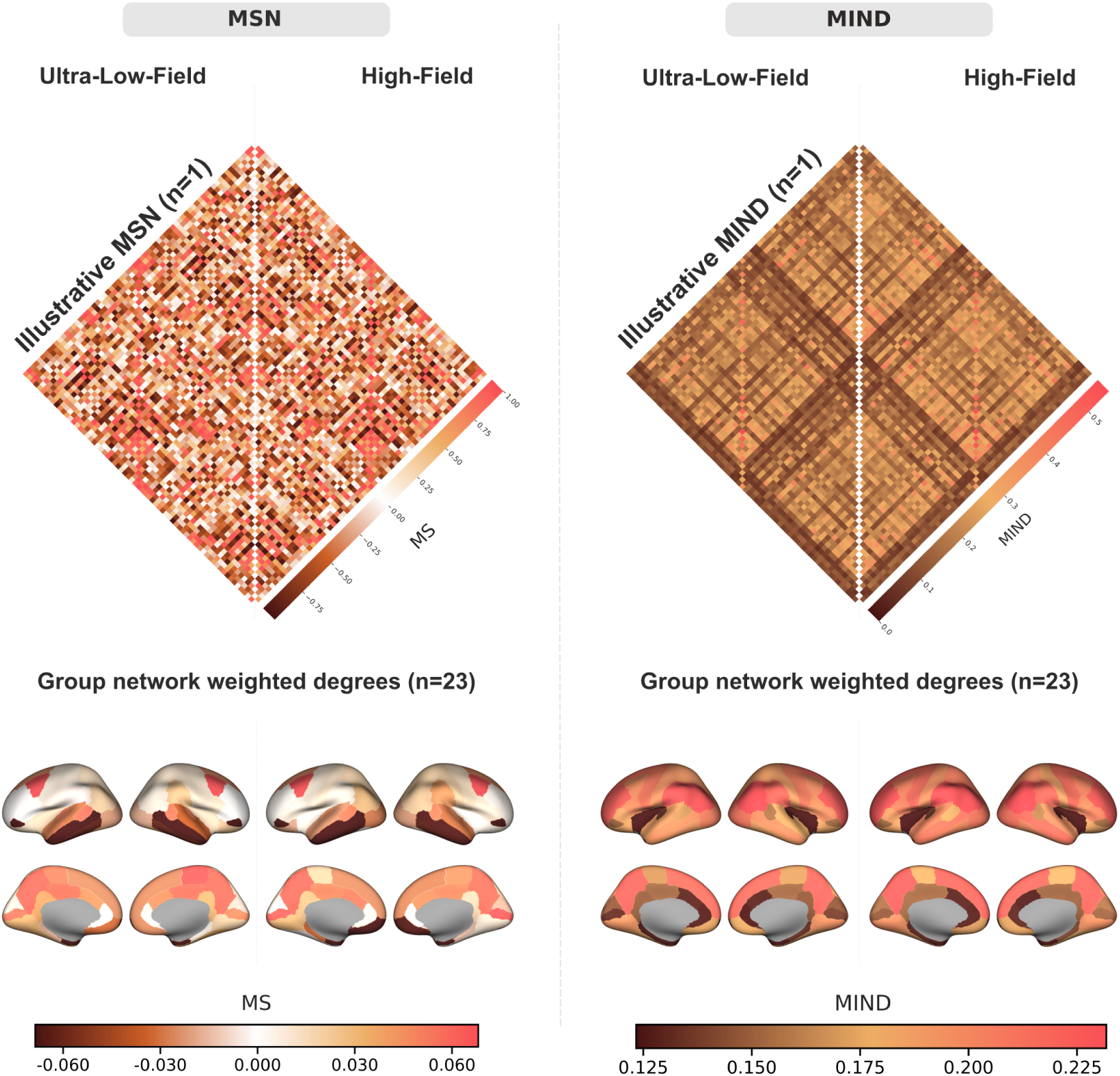
An illustrative network from the same randomly sampled participant in the primary cohort, and cortical surface maps of group network weighted degrees from high-field and ultra-low-field scans, per network type. **MS**: morphometric similarity. **MIND**: Morphometric INverse Divergence

### 2.5 Network Analyses

We constructed MSN and MIND individual structural networks for every subject, mesh resolution, and parcellation at HF and ULF. Our analyses were designed to answer the following questions:

1. **Cross-field agreement:** How similar are individual networks between HF and ULF, and does this scanner agreement differ across pre-processing methods and network types?
2. **Subject identifiability:** To what extent can an individual’s (MSN or MIND) network be matched to their own network across scans acquired at different field strengths (e.g., 3T vs 64mT) and across distinct scanners of the same field strength (e.g., two 64mT systems)?
3. **Effects of parcellation granularity:** How do parcellation choices (mesh resolution, atlas) affect agreement and identifiability measures?

Primary analyses for the first and second aim were performed in *fsnative* space using the Desikan-Killiany (DK) atlas and the five-feature set. This combination was selected a priori to balance high-resolution vertex-wise sampling with a relatively coarse yet robust 68-region atlas. Alternative cortical parcellations and mesh resolutions were used to assess robustness. Analyses compared pairs of scanners: HF–ULF_S1_ and ULF_S1_–ULF_S2_. When looking at the HF and ULF pairs, HF is considered the “ground truth”, as the structural connectomic methods have been widely applied and validated in HF images. The ULF scanner pair (ULF_S1_–ULF_S2_) is important to assess scanner effects and how stable ULF scanners are when extracting high-dimensional computational measures. Given that ULF_S1_ took place on the same day as the HF session, we focused on agreement in the HF–ULF_S1_ pair only. For the replication dataset, analyses included two pairs of HF – ULF (HF_1.5T_ – ULF and HF_3T_ – ULF) to assess cross-field agreement. We also analysed the HF_1.5T_ – HF_3T_ pair to compare MSN and MIND across high field strengths and to establish a benchmark.

We assessed cross-field agreement and ULF reproducibility of network edge weights at the global and node level (separately), by computing the Intraclass Correlation Coefficient, referred to as ICC(3,1) (Chen et al., 2018), and Lin’s Concordance Correlation Coefficient (CCC). Global values were calculated by vectorising the upper triangles of scanner-paired network matrices, and quantifying ICC and CCC per subject. These global values were then averaged across subjects to obtain a mean ± IQR for each scanner pair, reflecting overall linear similarity and concordance. For each metric, we evaluated super-resolution-method differences across networks, atlases, and scanner pairs: within each reliability metric (CCC and ICC3), two-sided paired Wilcoxon signed-rank tests comparing MRR and Supersynth were performed separately by network, atlas, and scanner pair. Raw p-values from the eight planned comparisons per metric were adjusted using the Benjamini–Hochberg FDR procedure. The same procedure was applied to evaluate network differences within super-resolution methods across atlases and scanner pairs. Node values were quantified using the same three statistics between rows of scanner-paired network matrices per subject. Similarly to global values, nodal values were averaged across subjects for each node. To assess network sensitivity to individual differences, we computed the subject-level identifiability metric (Amico and Goñi, 2018), which quantifies the extent to which within-subject similarity exceeds between-subject similarity. We computed Pearson correlations between vectorised HF-ULF network pairs for all subjects. Specifically, we vectorised the upper triangle of each subject’s adjacency matrix (edge-weight MSN or MIND matrix) and correlated HF and ULF vectors (as well as ULF-ULF vectors for reproducibility) within and between subjects, yielding an N x N correlation matrix. We then averaged the diagonal elements quantifying within-subject similarity to obtain I_self_, and the off-diagonal elements quantifying between-subject similarity to obtain I_others_, and finally calculated the differential identifiability as:

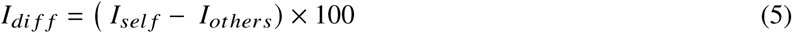

To assess the effects of mesh resolution on agreement and identifiability in morphometric networks, we reconstructed subjects’ MIND networks using a coarser *fsaverage5* mesh compared to the *fsnative* mesh, and computed CCC and I_diff_ for the new networks. Because MSN networks are based on regional summary statistics and are expected to be relatively insensitive to mesh resolution, we did not reconstruct MSNs on the lower-resolution mesh.

## 3 Results

### 3.1 Agreement and reproducibility at the edge-level

For each scanner pair and subject, we computed edgewise Lin’s CCC and ICC3, and summarised these values (mean [IQR] across subjects) to obtain network-level agreement for both MSN and MIND. Because CCC captures both correlation and bias, we focus on reporting CCC in the main text, while ICC is reported in Supplementary Material.

For brevity, we use the term **cross-field agreement** to refer to agreement between HF and ULF scans (HF–ULF), **reproducibility** to refer to agreement between repeated ULF scans (ULF–ULF), and **cross-scanner agreement** to refer to correspondence between HF scanners of different field strengths (HF_1.5T_-HF_3T_).

Overall, for both super-resolution methods, ULF MIND networks demonstrated stable edge-level cross-field agreement 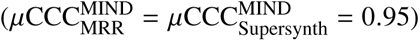 and ULF reproducibility (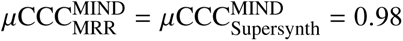; see Table 2 for full details of the MSN comparisons). MSNs also showed strong, but slightly lower, cross-field agreement with a higher IQR, as shown in Figure 4 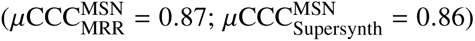, as well as high ULF reproducibility 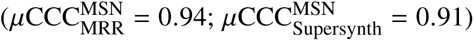.

**Figure 4.**
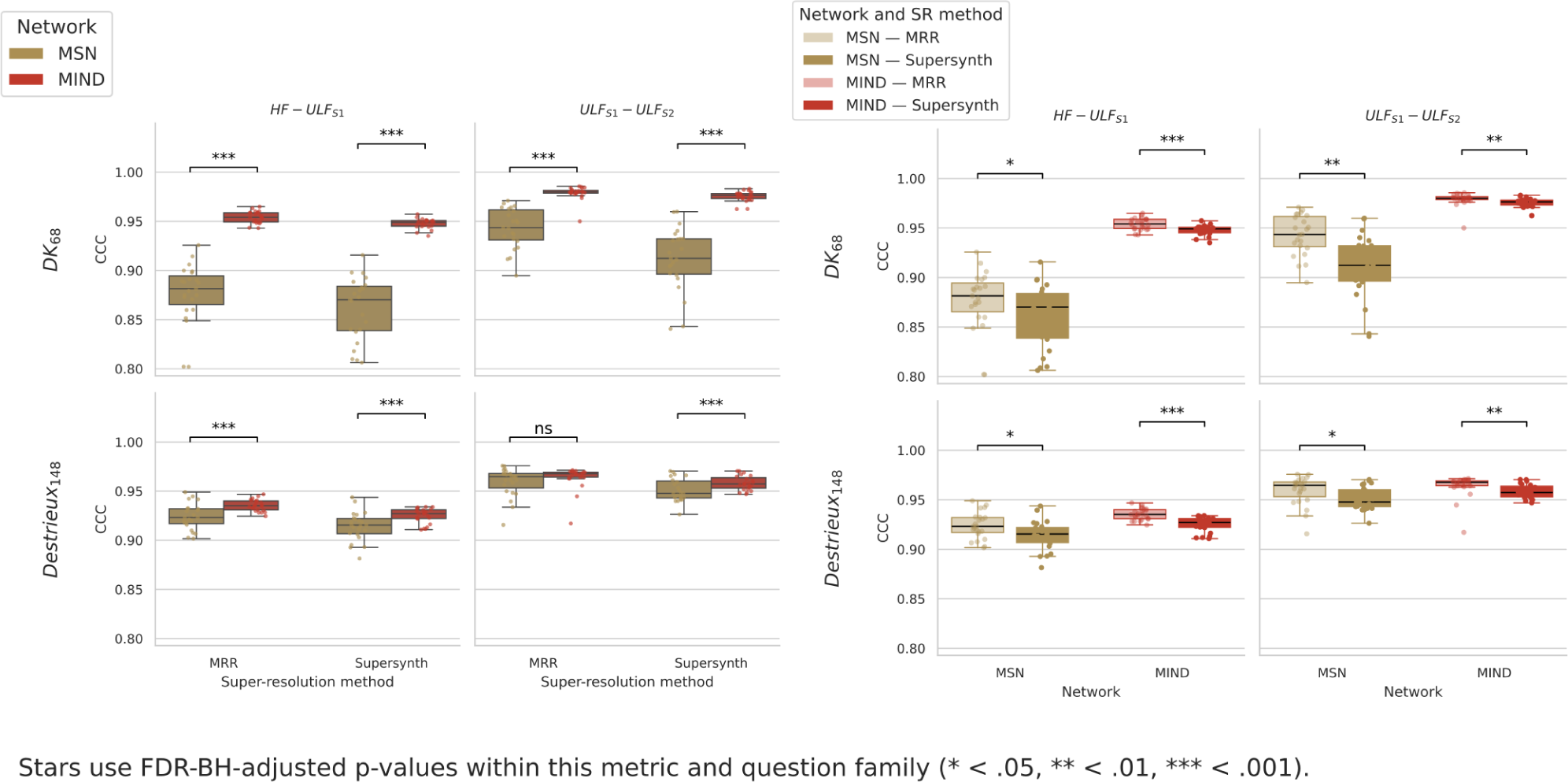
Edge-level CCC across network types, super-resolution methods, atlases, and scanner pairs. The vertical axis displays CCC values; the horizontal axis shows either the the super-resolution method (left) or network type (right). Centre lines represent the medians, box limits indicate the 25th and 75th percentiles, and whiskers extend to the most extreme data points within 1.5 x IQR. Individual points represent values from individual participants. Both panels display the same underlying data, but arranged differently for readability: **(Left)** MSN versus MIND comparison within each super-resolution method **(Right)** MRR versus Supersynth comparison within each network type.

**Table 1.** Primary and replication cohort demographics and their corresponding MRI scanning sessions. *Note.* 64mT S1 and 64mT S2 denote two distinct 64mT systems with identical hardware and software, in different locations. Of the 19 participants in the replication cohort, one participant was missing a 1.5T scan.

| Cohort | N | Age (years) | Sex (% F) | Scanning Sessions |
| --- | --- | --- | --- | --- |
| **Primary Cohort** | 23 | 20–69 | 48% | 1 HF<sub>3T</sub><br>1 ULF<sub>64mT</sub> (S1)<br>1 ULF<sub>64mT</sub> (S2) |
| **Replication Cohort** | 19 | 24–49 | 63% | 1 HF<sub>3T</sub><br>1 HF<sub>1.5T</sub><br>1 ULF<sub>64mT</sub> |

**Table 2.** Summary of between-scanner edge-level brain network agreement for MSN and MIND networks. CCC values are reported as mean [IQR] across 2,278 edges for the DK atlas and 10,878 edges for the Destrieux atlas, per subject. Maximum CCC values within a category (agreement or reproducibility) are highlighted in bold.

| Atlas | SR method | HF–ULF<sub>S1</sub><br>Cross-field agreement |  | ULF<sub>S1</sub>–ULF<sub>S2</sub><br>Reproducibility |  |
| --- | --- | --- | --- | --- | --- |
| | | $\mu\text{CCC}_{\text{edge}}^{\text{MSN}}$ | $\mu\text{CCC}_{\text{edge}}^{\text{MIND}}$ | $\mu\text{CCC}_{\text{edge}}^{\text{MSN}}$ | $\mu\text{CCC}_{\text{edge}}^{\text{MIND}}$ |
| DK<sub>68</sub> | MRR | 0.87 [0.03] | **0.95 [0.009]** | 0.94 [0.03] | **0.98 [0.002]** |
|  | Supersynth | 0.86 [0.04] | **0.95 [0.007]** | 0.91 [0.04] | **0.98 [0.005]** |
| Destrieux<sub>148</sub> | MRR | 0.92 [0.02] | **0.94 [0.01]** | 0.96 [0.01] | 0.96 [0.01] |
|  | Supersynth | 0.91 [0.02] | **0.92 [0.01]** | 0.90 [0.02] | **0.96 [0.01]** |

Edge-level agreement and reproducibility scores of MIND networks outperformed MSN scores, as shown in Figure 4. Coarseness of the cortical parcellation affected agreement and reproducibility differently for MSN and MIND. Using the DK atlas showed higher CCC values across pairs of scanners for MIND (in both super-resolution methods), whereas the Destrieux atlas resulted in higher agreement and reproducibility in MSNs. MIND network correspondence also displayed higher CCC values and narrower IQRs than MSN, indicating less variability in edge-wise agreement across subjects. IQRs in MSNs were narrower in the Destrieux space compared to DK.

### 3.2 Agreement and reproducibility at the node level

We also assessed regional patterns of agreement by computing node-wise CCC for MSN and MIND networks and averaged across subjects, to obtain *μ*CCC_region_ per super-resolution method and scanner pair. We found comparable scanner agreement patterns when using MRR or Supersynth T2w input (Figure 5). Regional CCC maps were highly similar between the two methods, indicating that the choice of super-resolution method has minimal impact on regional reliability. Lower scanner agreement was most apparent in orbitofrontal regions, whereas primary sensory motor areas showed consistently high agreement.

**Figure 5.**
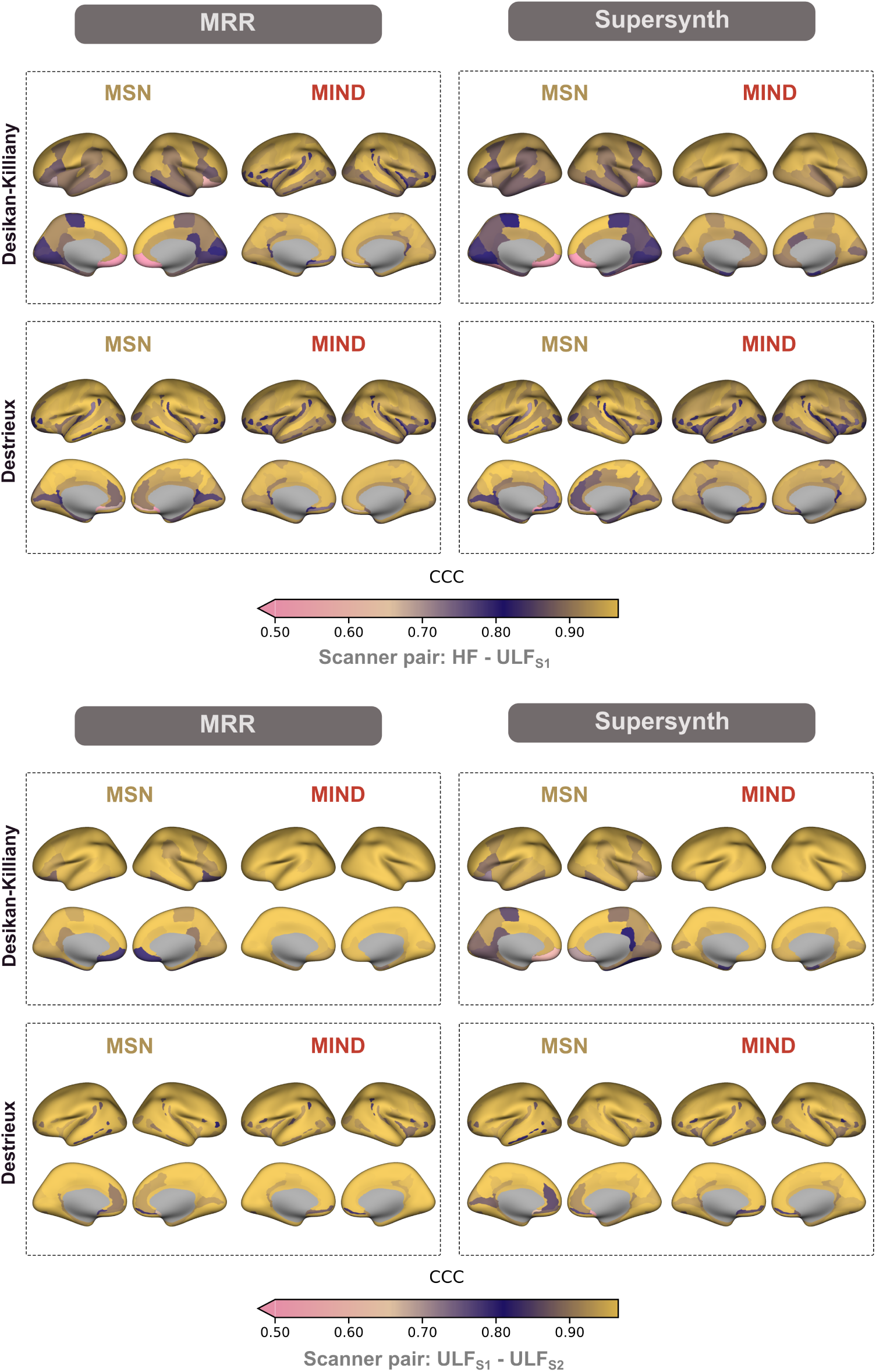
Scanner agreement at the node level for MSN and MIND networks across input types captured by CCC. **(Top)** Scanner pair: HF-ULF_S1_ **(Bottom)** Scanner pair: ULF_S1_ - ULF_S2_. Colour scale lower bound was set to 0.5; values below this bound are shown at the minimum colour.

#### Replication Cohort: cross-field and cross-scanner agreement

After applying the same pre-processing and analysis steps developed in the primary cohort on the replication cohort, our findings show highly similar patterns, notably high HF-ULF scanner agreement ( see Table 3). We also note that HF-ULF agreement measures are higher in MIND networks compared to MSNs in both pairs (1.5T and 3T), as shown in the primary cohort. In this cohort, Supersynth yielded better agreement measures than MRR, unlike in the primary cohort.

**Table 3.** Summary of between-scanner edge-level brain network agreement for MSN and MIND networks in the replication cohort. CCC values are reported as mean [IQR] across 2,278 edges for the DK atlas and 10,878 edges for Destrieux atlas, per subject. Results are reported in the DK_68_ cortical atlas space. Maximum CCC values between MSN and MIND within a category (agreement or reproducibility) are highlighted in bold.

| Atlas | SR method | HF 1.5T–ULF<br>Cross-field agreement |  | HF 3T–ULF<br>Cross-field agreement |  |
| --- | --- | --- | --- | --- | --- |
| | | $\mu\text{CCC}_{\text{edge}}^{\text{MSN}}$ | $\mu\text{CCC}_{\text{edge}}^{\text{MIND}}$ | $\mu\text{CCC}_{\text{edge}}^{\text{MSN}}$ | $\mu\text{CCC}_{\text{edge}}^{\text{MIND}}$ |
| DK<sub>68</sub> | MRR | 0.81 [0.05] | **0.94 [0.02]** | 0.81 [0.04] | **0.94 [0.02]** |
|  | Supersynth | 0.86 [0.04] | **0.96 [0.01]** | 0.85 [0.06] | **0.96 [0.01]** |
| Destrieux<sub>148</sub> | MRR | 0.89 [0.03] | **0.92 [0.02]** | 0.90 [0.02] | **0.92 [0.02]** |
|  | Supersynth | 0.93 [0.01] | **0.94 [0.01]** | 0.93 [0.01] | **0.94 [0.02]** |

For HF scanner-agreement in the 1.5T – 3T pair, see Supplementary Table S1. CCC measures were high for both networks, with MIND slightly outperforming MSN in the DK and Destrieux atlas space.

### 3.3 Subject identifiability in MSN and MIND networks

To assess whether subjects can be identified across scanners, we computed the identifiability measure proposed by (Amico and Goñi, 2018). Figure 6 illustrates the identifiability matrices, where diagonal values denote within-subject correlations, and off-diagonal values represent between-subject correlations.

**Figure 6.**
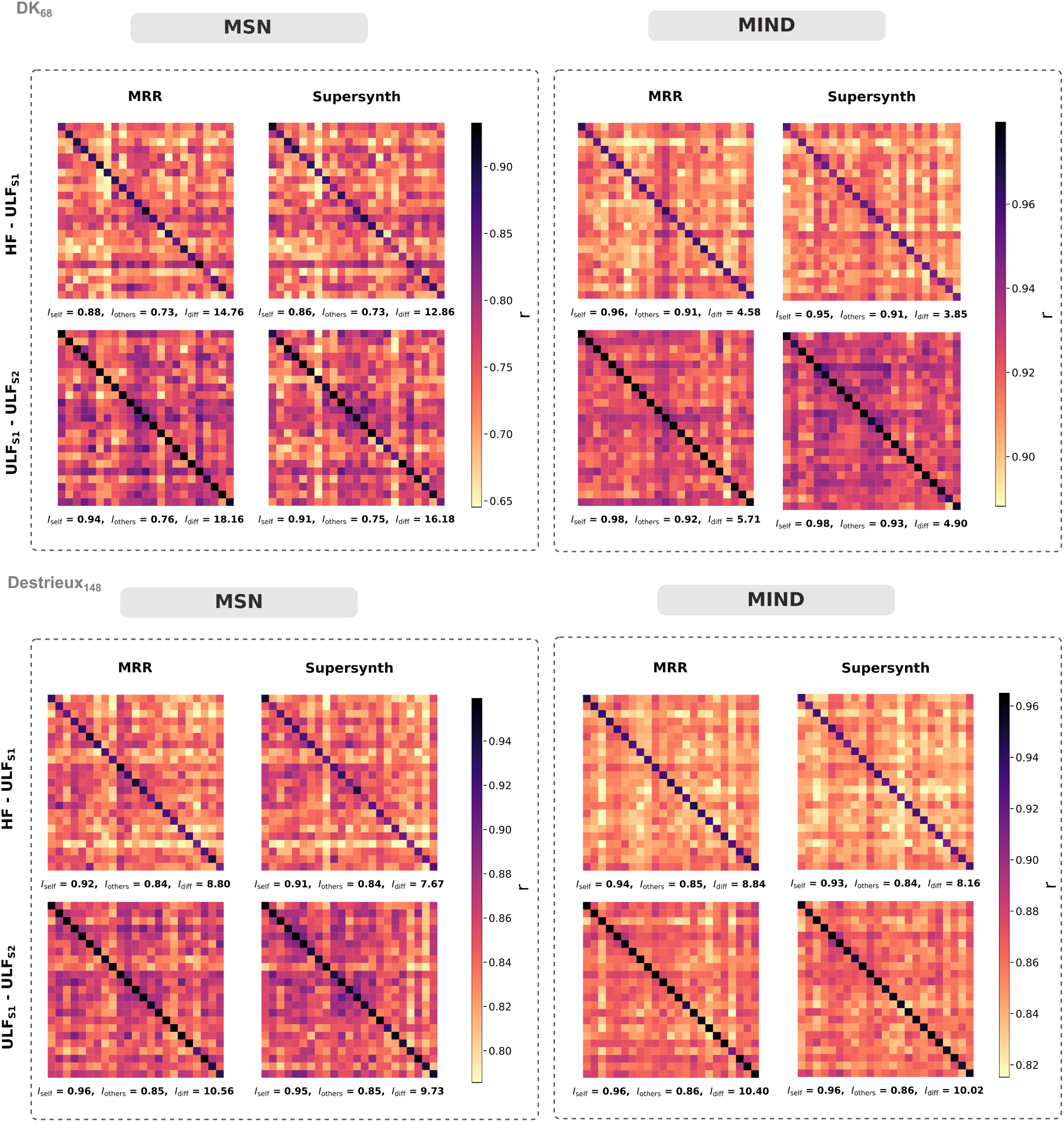
MSN and MIND identifiability plots showing subject-level differentiation across scanners per SR method and cortical atlas. **(Top)** MSN and MIND identifiability plots in the DK_68_ cortical atlas. **(Bottom)** MSN and MIND identifiability plots in the Destrieux_148_ cortical atlas.

Within the ULF pair (ULF_S1_ – ULF_S1_), the subject identifiability diagonal was visually distinct for both networks. Within the HF and ULF pair, I_self_ remained consistently higher than I_others_ for all scanner pairs in both networks, indicating that subjects were still best matched to themselves, although the difference was reduced compared to within ULF-identifiability (see Table 4 for full comparisons). The higher I_diff_ values for MSNs are driven by a much larger distinction between within- and between-subject similarity than for MIND. For MSNs, _self_ exceeded _others_ by at least 0.13, whereas for MIND this difference was at most 0.05.

**Table 4.** Summary of subject network identifiability for MSN and MIND, across scanners, atlases, and super-resolution methods. Maximum identifiability values within each network are highlighted in bold.

| Atlas | SR method | MSN |  | MIND |  |
| --- | --- | --- | --- | --- | --- |
| | | $I_{\text{diff}} \text{ (HF-ULF)}$ | $I_{\text{diff}} \text{ (ULF-ULF)}$ | $I_{\text{diff}} \text{ (HF-ULF)}$ | $I_{\text{diff}} \text{ (ULF-ULF)}$ |
| **DK<sub>68</sub>** | **MRR** | 14.76 | **18.19** | 4.58 | **5.71** |
|  | **Supersynth** | 12.86 | **16.18** | 3.85 | **4.90** |
| **Destrieux<sub>148</sub>** | **MRR** | 8.80 | **10.56** | 8.84 | **10.40** |
|  | **Supersynth** | 7.67 | **9.73** | 8.16 | **10.02** |

Subjects were better identified by their networks across field strengths and scanners when using the Destrieux_148_ atlas than the DK_68_. When comparing identifiability in the DK_68_ across super-resolution methods, MRR yielded higher differential identifiability than Supersynth, with an increase of Δ*I*_diff_ = *I*_diff_^MRR^ − *I*_diff_^Supersynth^ = 1.9 percentage points for MSNs and 0.73 for MIND networks. Similar differences were observed for the Destrieux_148_ atlas.

#### Replication Cohort: subject identifiability

Results were replicated in the independent cohort (Figure 7). Subjects’ brain networks were identified across field-strengths in the 3T – 64mT and 1.5T – 64mT scanner pairs, with MSNs having higher I_diff_ values than MIND, as found in the primary cohort. We again observed a larger separation between within- and between-subject similarity in MSN than in MIND. For MSNs, I_self_ exceeds I_others_ by at least 0.13, whereas for MIND this difference is at most 0.04. As participants in this cohort had two HF scans, we also analysed data from the HF scanner pair (1.5T – 3T) scanner pair, as a benchmark to assess ULF results. The difference in identifiability metrics between MSN and MIND in the HF_1.5T_ – HF_3T_ pair was comparable to the difference observed in the HF-ULF scanner pairs. For MSNs, I_self_ exceeded I_others_ by 0.23, whereas for MIND this difference is 0.07.

**Figure 7.**
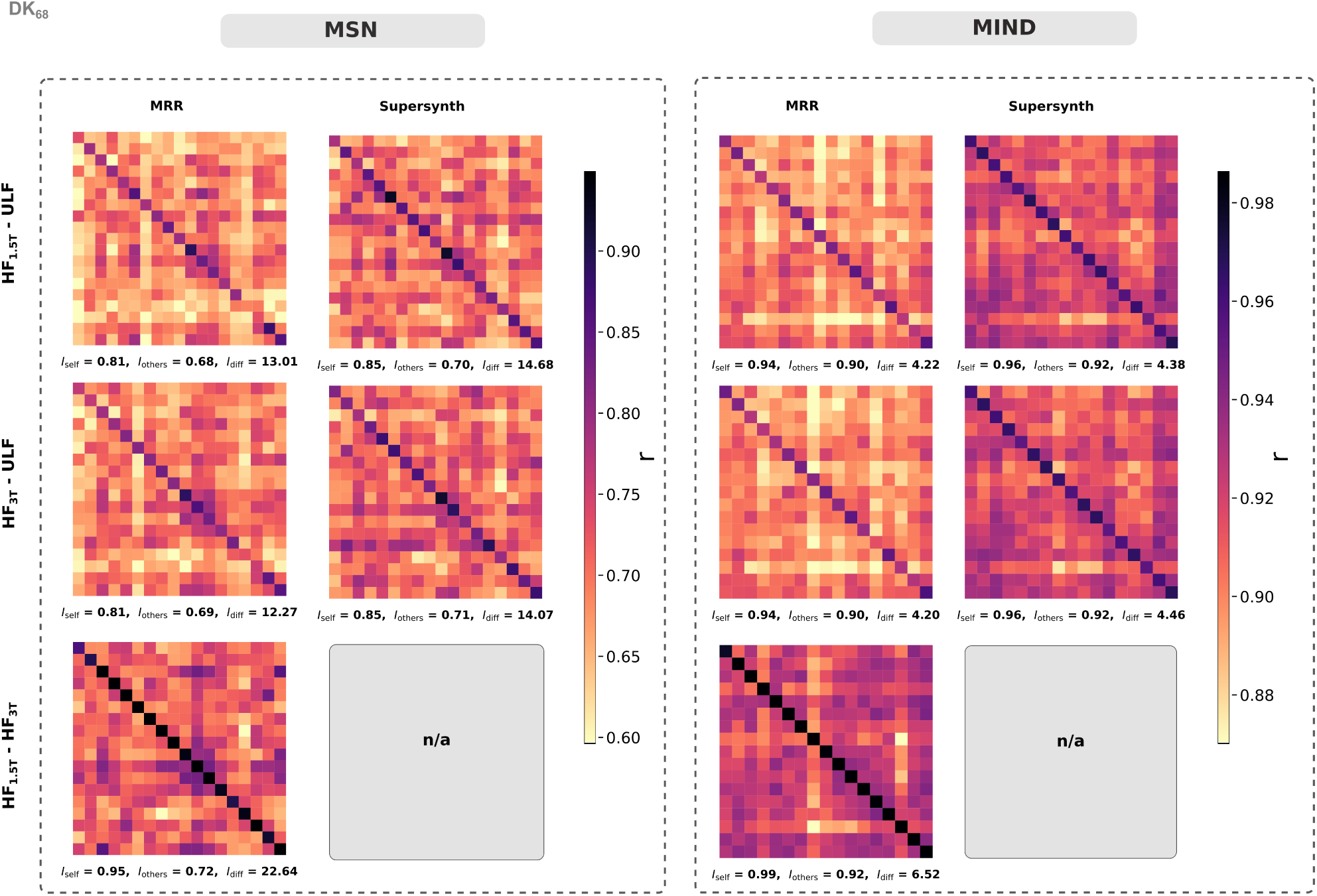
MSN and MIND identifiability plots showing subject-level differentiation across scanners (1.5T and 3T HF, and 64mT ULF) in the replication cohort. Results reported in the DK_68_ cortical atlas space.

### 3.4 Network granularity effects on agreement and identifiability

We repeated the global-level and node-level analyses previously described on the newly constructed MIND networks. Agreement between scanners was systematically higher when networks were reconstructed in *fsnative* space than in *fsaverage5* across both scanner pairs and atlases (all p ≤ 0.001; Figure 8). See Supplementary Table S2 for full edge-level CCC comparisons between meshes. Differences between super-resolution methods across meshes were smaller: MRR yielded equal or slightly higher CCC than Supersynth, with significant effects mainly in the *fsnative* space, and largely comparable performance in the *fsaverage5* space.

**Figure 8.**
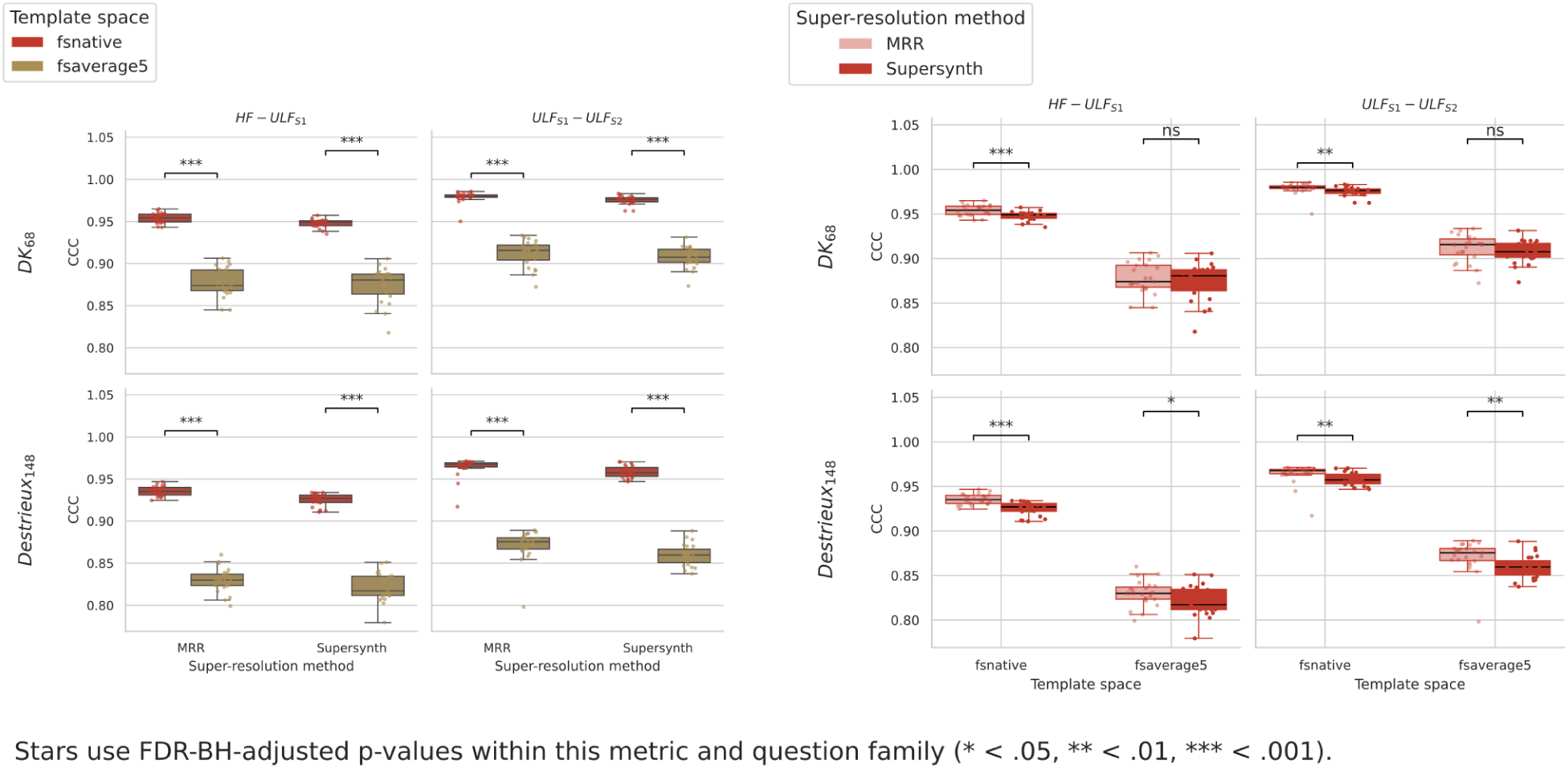
Edge-level CCC across mesh resolutions, atlases and scanner pairs. The vertical axis displays CCC values; the horizontal axis shows either the the super-resolution method (left) or mesh (right). Centre lines represent the medians, box limits indicate the 25th and 75th percentiles, and whiskers extend to the most extreme data points within 1.5 x IQR. Individual points represent values from individual participants. Both panels display the same underlying data, but arranged differently for readability: **(Left)** *fsnative* versus *fsaverage5* within each super-resolution method **(Right)** MRR versus Supersynth within each mesh.

For MIND networks, we next examined regional scanner agreement as a function of template mesh. Across both atlases and super-resolution methods, reconstruction in *fsnative* space yielded higher node-wise CCC than in *fsaverage5*, but the spatial pattern of agreement was similar between meshes (Figure 9). In both cases, the lowest CCC values were observed in orbitofrontal and temporal pole regions, whereas primary sensorimotor and occipital cortices showed consistently high agreement.

**Figure 9.**
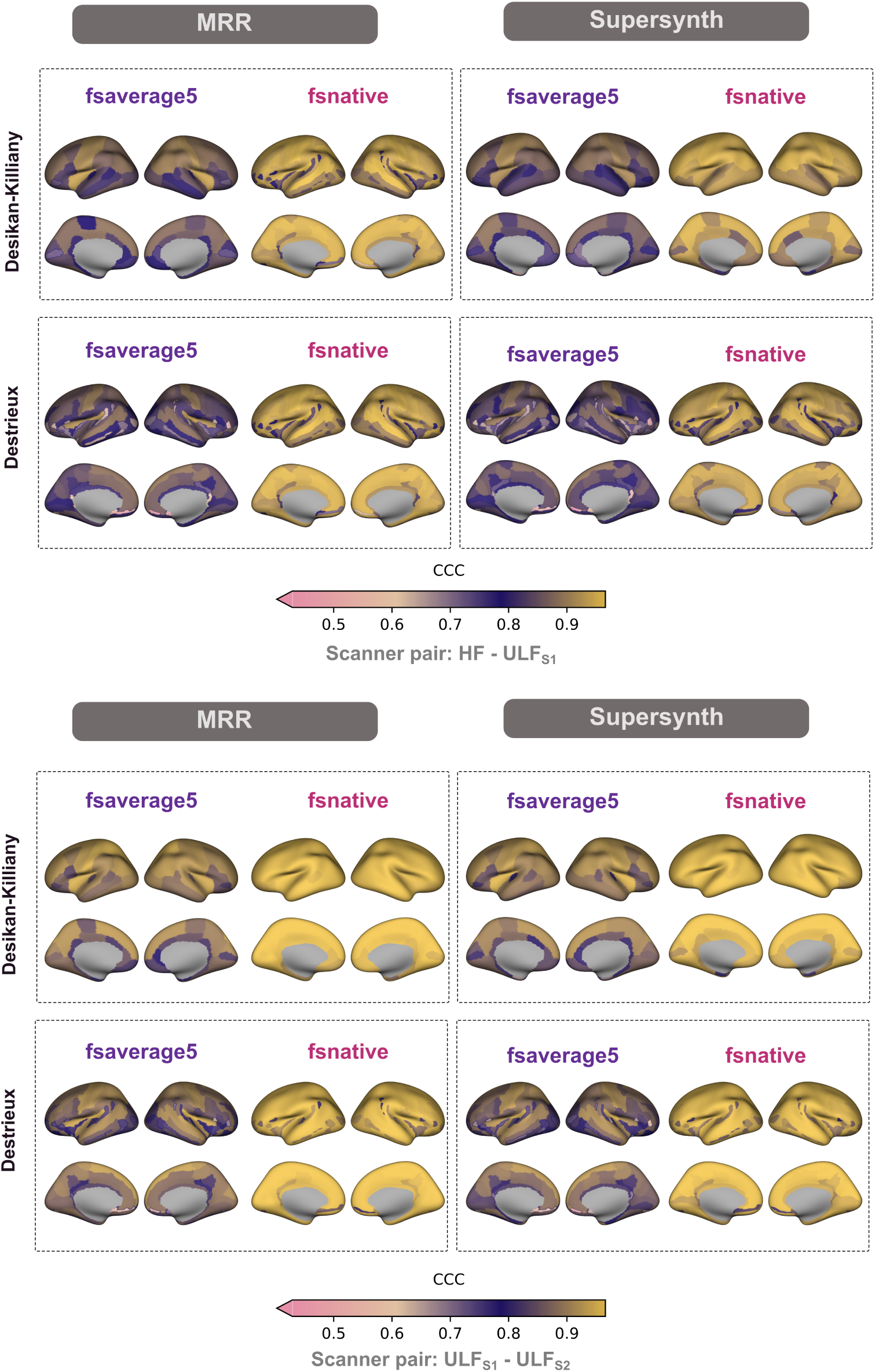
Scanner agreement at the node level for MIND networks across meshes and SR methods captured by CCC. **(Top)** Scanner pair: HF-ULF_S1_ (Bottom) Scanner pair: ULF_S1_ - ULF_S2_. Colour scale lower bound was set to 0.5; values below this bound are shown at the minimum colour.

**Figure 10.**
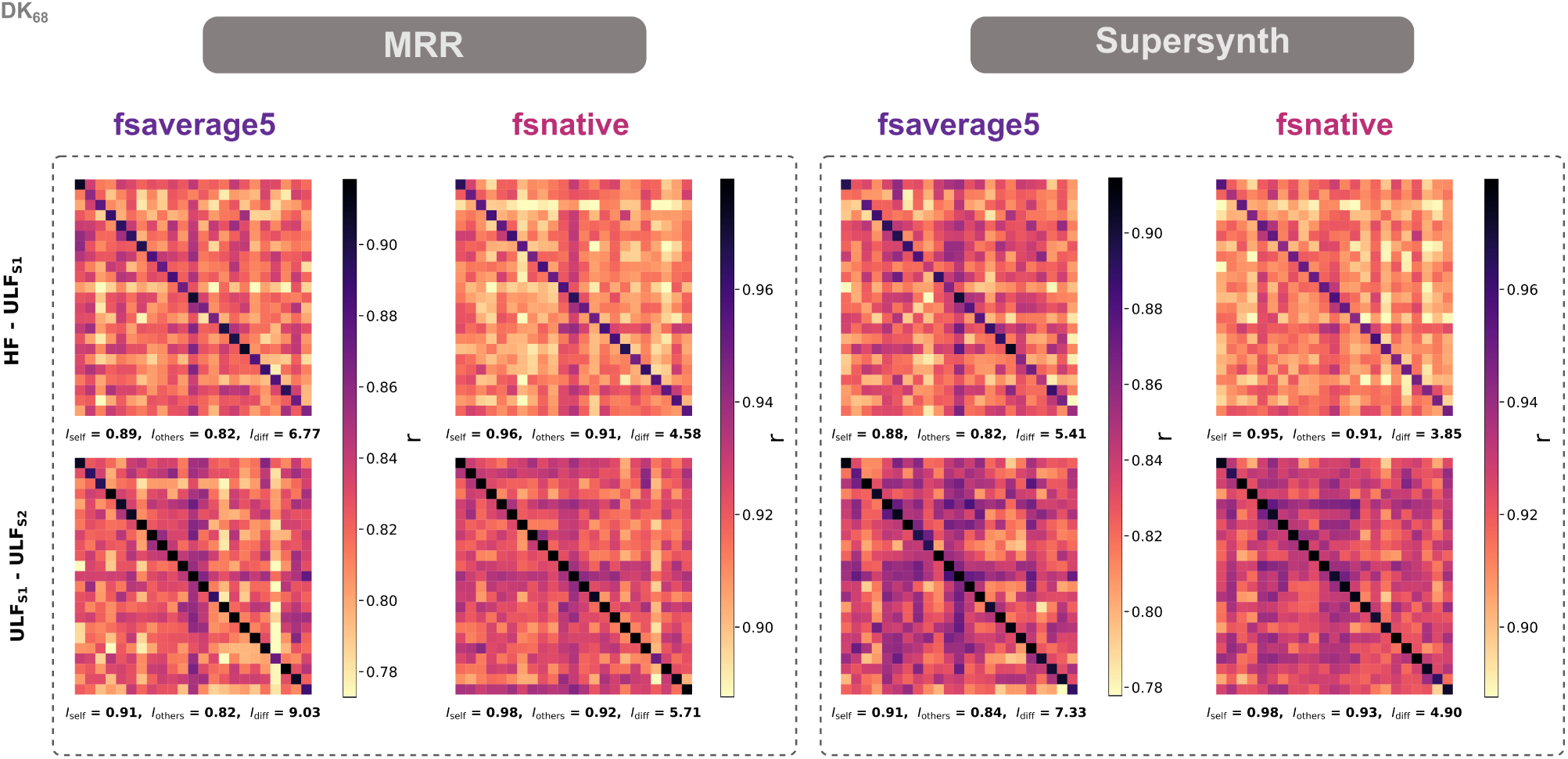
Identifiability plots showing MIND network subject-level differentiation across scanners using different mesh resolutions. Results reported using DK_68_ cortical atlas.

In contrast, MIND networks constructed using the coarser mesh showed higher identifiability measures in both the DK_68_ (Figure 9) and the Destrieux_148_ atlas. In both super-resolution methods, the difference ranged between Δ*I*_diff_ = *I*_diff_^fsaverage5^ − *I*_diff_^fsnative^ = 1.56 and 2.19 percentage points for the HF–ULFS1 pair, and between 2.43 and 3.32 for the ULF_S1_–ULF_S2_ pair. Similar differences were observed in the Destrieux_148_ atlas (Supplementary Figure S2)).

## Discussion

In this work, we assessed the feasibility of constructing morphometric networks in a set of 23 healthy adult subjects using paired high and ultra-low-field MRI data, and compared two different approaches in brain network construction. We first segmented structural T2w scans acquired with a GE 3T, as well as two Hyperfine Swoop ® 64mT scanners. After super-resolving the ULF scans with 2 different methods, we constructed MSN and MIND networks using the FreeSurfer recon-any tool for every mesh and cortical atlas combination. Agreement between HF and ULF was highest for MIND networks compared to MSN in both DK and Destrieux-based networks, as was the ULF reproducibility. Subjects were highly identifiable by their brain network across field-strengths and same-strength scanners, with a high subject differential identifiability using MSNs. In additional analyses, and to compensate for the small sample size in the primary cohort, we showcased the generalisability of the results in an independent cohort of 19 healthy adults scanned on two HF scanners (1.5T and 3T) and one Hyperfine Swoop ® 64mT scanner, by assessing HF-ULF agreeement in the 3T–64mT and 1.5T–64mT scanner pairs, as well as cross-HF agreement in the 1.5T–3T pair.

This study built on previous work by Váša et al., 2025 on this cohort showing high HF-ULF correspondence and ULF test-rest reliability in brain volume estimates. We found that structural similarity networks can be reliably constructed from ULF scans using T2w images, with high 3T correspondence and excellent between-ULF-scanner reliability. Test–retest reliability of MSN and MIND networks in the high-field replication cohort provided a benchmark for interpreting the ULF findings. Differences observed between MSN and MIND in the 1.5T–3T pair were similar to those observed in other pairs (HF–ULF and ULF–ULF). This therefore suggests that the effects we see in HF–ULF and ULF–ULF comparisons are not solely attributable to ULF-specific artefacts.

To our knowledge, this is the first study in its kind to investigate the application of analytical approaches of brain connectomics in the context of ULF MRI. In our primary dataset, T2w images were not acquired in the second ULF session. To obtain isotropic T2w-like contrast for those scans, we used the corresponding ULF T1w images as input to Supersynth to generate synthetic iso-T2w images. Although Váša et al., 2025 reported lower reliability and correspondence for morphometric measures in T1w than in T2w scans, estimates of agreement found in this work remained high. Additionally, since the original study on this dataset investigated the impact of contrast on reported regional brain volumes, this work primarily focused on differences between network types, cortical parcellations, and mesh resolutions. However, scans obtained after multi-resolution-registration (MRR) of three individual anisotropic T2w acquisitions showed better cross-scanner agreement, reproducibility, and subject identifiability, compared to synthetic T2w scans generated with Supersynth using the subject’s corresponding T1w scan. One possible explanation is the previously mentioned issue with T1w scans, where their low correspondence may propagate into the synthetically generated T2w. Another explanation could be that the Supersynth model, trained primarily on adults, may bias reconstruction anatomy towards the training distribution, resulting in a slight compression of individual differences. In their review of construction choices on structural brain network measures, (Qi et al., 2015) stress that network measures are highly sensitive to preprocessing steps. This major super-resolution step in the pipeline could be at the root of the drop in agreement. Interestingly, we saw this pattern however was reversed in the replication cohort. This discrepancy may be related to lower quality scans in the replication cohort compared to the primary cohort. This suggests that in lower quality acquisitions, a super-resolution model could be used to effectively denoise and regularise morphometry to improve between-scanner agreement, while still preserving network-level morphometric features. That said, applying super-resolution models can come at the expense of diminishing individual differences and accuracy in predictive analyses. Furthermore, applying a model that was trained on adults to paediatric or atypical cohorts (e.g., clinical) could systematically alter morphometric distributions, resulting in even lower agreement metrics.

MIND networks outperformed MSNs at capturing nodal relationships across field strengths, showing systematically higher regional agreement values and less variability across cortical regions. Conversely, MSN-based nodal agreement was more heterogeneous. One possible explanation is that higher-resolution vertex data in MIND reduce averaging artefacts contrary to MSN’s regional estimates. However, reduced variance (and subject identifiability) could also lead to loss of biological relevance. Agreement in MSN improved and stabilised when using the finer cortical Destrieux parcellation, showing that even a moderate increase in parcellation resolution can result in more anatomically coherent nodes and better network stability.

Our findings also showed that subjects can be identified by their brain networks across scanners and field strengths using both super-resolution methods, though as mentioned, MRR-based input resulted in higher agreement compared to Supersynth. This is further evidence of the reliability of ULF scans, their stability across sessions, as well as evidence for both networks’ idiosyncrasy. Compared to MIND, each subject’s MSN was more identifiable across scanning sessions and field-strengths. As previously mentioned, the low variance in between-scanner agreement in MIND networks can be a sign of robustness, or bias that masks subject specificity, leading to lower subject identifiability. MSN networks produced a clearer gap between within-subject and between-subject similarity than MIND networks, which directly explains their higher differential identifiability. Within-subject similarity improved for MSN when using a coarser atlas, and a finer atlas for MIND. This is in line with with the predictions shown in the recent methodological work on MIND in a normative dataset from the UK Biobank cohort by Davies et al., 2026, who found that although finer parcellations show higher edge-level variability, they can be more sensitive to between-subject differences, indexed by higher sex-classification accuracy. While we do not perform predictive modelling in this study, differential identifiability similarly quantifies how distinct individual network patterns are across subjects, and thus relates to the potential of these networks to support subject-level prediction. This opens a promising avenue for more work using brain connectomics in ULF such as looking at the predictive power of brain networks and their associations to behavioural outcomes (e.g., cognitive or clinical measures). While Amico and Goñi, 2018 developed the differential identifiability framework in the context of functional MRI and Principal Component Analysis reconstruction, the original paper and later work explicitly mention the potential for extending it to structural connectivity as a natural next step. In this work, we extended this approach to proxy measures of structural connectivity, and demonstrated its feasibility and robustness in ULF MRI.

Our analyses looking at the influence of cortical parcellation and mesh resolution in MIND networks showed that agreement and reliability were highest when using the native high-resolution mesh, compared to the coarser *fsaverage5* mesh. These findings are again consistent with those by Davies et al., 2026, where authors show that changing vertex mesh resolutions via interpolation can bias similarity estimates. They showed that the lowest-resolution template performed worse in terms of correspondence with gene co-expression, and advised against performing additional mesh down-sampling in order to preserve biological validity and reduce edge-level noise. This recommendation may be particularly relevant for lower-resolution scans. Additionally, the authors demonstrated that parcellation granularity and region size systematically influence MIND estimates, with smaller parcels in finer atlases showing higher edge-level variability, and coarser atlases showing stronger correspondence with gene co-expression. In our data, MIND networks constructed with the coarser Desikan-Killiany atlas showed higher scanner agreement than those using the finer Destrieux atlas, suggesting that there may be an upper limit to consider practically when processing ULF data, to ensure stable MIND network estimation.

While finer cortical parcellations decreased agreement measures in MIND, they increased subject identifiability. This aligns with network neuroscience literature showing that individual specificity or “fingerprinting” of functional and structural connectivity generally improves with finer parcellations (Jiang et al., 2023; Zimmermann et al., 2018; Finn et al., 2015). In contrast, brain network identifiability was higher when using a coarser mesh. This was the case between HF-ULF and ULF-ULF scanners in both the primary and replication cohort, as well as the HF scanner pair (HF_1.5T_-HF_3T_) in the replication cohort. One plausible interpretation is that the coarser mesh may effectively average vertex-level noise and small misregistration errors, improving the stability of subject-specific morphometric patterns across scanners, whereas the high-resolution mesh could be more sensitive to scanner-specific variability, at the cost of lower cross-scanner identifiability. Taken together, these findings indicate that, in healthy adults, cross-scanner agreement is best achieved with MIND networks using a coarser cortical atlas on the native mesh, whereas subject identifiability is maximised with MSN networks using a finer cortical atlas. Overall, our results show that we can reliably build structural similarity networks using ULF data in a cohort of healthy adults. We reconstructed the network by using a combination of cortical atlases and mesh resolutions. We found that optimal parcellation scale depends both on the similarity network (MIND versus MSN) and on the target measure (cross-scanner stability versus within-subject identifiability), echoing prior work on resolution trade-offs in structural networks (Zalesky et al., 2010; Romero-Garcia et al., 2012; Davies et al., 2026).

### Limitations and Future Work

While we successfully replicated our results, the sample sizes of both the primary and the replication cohort were small (N = 23 and N = 19 respectively). There is still the question of whether this would stand in a bigger cohort with more noise in the data, data of lower quality, or data acquired on other software versions of the Hyperfine Swoop ® scanner (Stockbridge et al., 2026). As scanner software and reconstruction models are updated, and as offline super-resolution algorithms evolve, we expect overall patterns of agreement and identifiability to remain stable in similar cohorts, if not improve with newer hardware and software iterations. However, minor differences in absolute estimates and robustness of results may be observed across software versions and reconstruction pipelines. It is important to note that our results pertain and generalise to healthy adults only. While similar high levels of agreement may not be achievable in developmental or clinical populations, these findings provide a useful benchmark and reference point for future ULF studies in more challenging cohorts. Hence, the next step would be to assess the reliability of brain networks in younger cohorts such as adolescents, young children, and infants, where artefact presence is higher in scans, tissue contrast is affected by rapid neurodevelopment, and brain segmentation quality and precision requires more developmentally tailored tools and rigorous quality control (Barkovich et al., 2019; Dubois et al., 2021). However, given that previous studies have already validated HF-ULF agreement in adolescents and young children at both regional and vertex-levels (Pretzsch et al., 2025; Cooper et al., 2024; Ringshaw et al., 2026), we hypothesise that the analysis pipeline presented here could be suitable for paediatric imaging, with minor modifications to fine-tune it for developmental cohorts. Additionally, the brain’s fingerprint will be an evolving signature in younger cohorts and clinical groups. Future work should examine how this individual network fingerprint evolves across the lifespan, and whether networks constructed at earlier timepoints can predict subsequent changes in the structural connectome, age-related reorganisation, and associated cognitive, behavioural, and pathological outcomes.

## Data and Code Availability

Imaging data from the primary cohort are openly available at: OpenNeuroDS006557. Code is available at: GithubRepo

## Author Contributions

**Hajer Karoui**: Conceptualisation, Formal analysis, Software, Methodology, Validation, Visualisation, Writing - original draft, Writing - review and editing. **Niall J. Bourke**: Resources, Software, Supervision, Writing - review and editing. **James H. Cole**: Data Collection (replication cohort), Data quality control, Writing - review and editing **Rui W. Yeow**: Data Collection (replication cohort), Data quality control, Writing - review and editing **Fred Dick**: Funding acquisition, Resources, Data Collection (replication cohort), Writing - review and editing **Raha Razin**: Data Collection (replication cohort) **David L. Thomas**: Data Collection (replication cohort), Writing - review and editing **Kirsten W. R. Schröder**: Data Collection (replication cohort) **David M. Cash**: Data Collection (replication cohort), Writing - review and editing **Martina Bocchetta**: Data Collection (replication cohort), Writing - review and editing **Steve C.R. Williams**: Funding acquisition, Resources, Supervision, Writing - review and editing. **František Váša**: Data Collection (primary cohort), Methodology, Resources, Supervision, Writing - review and editing.

## Declaration of the use of AI

AI was used for rewording and checking code used for analysis and visualisation. Models used include claude-sonnet-4-6, claude-sonnet-5, gpt-5.1, and gpt-5.6-terra.

## Appendix: Supplementary Materials

### Edge-level cross-HF-scanner agreement in the replication cohort

**Table S1.** Summary of edge-level HF cross-scanner agreement for MSN and MIND networks.

| Atlas | HF 1.5T–HF 3T<br>Cross-field agreement |  |
| --- | --- | --- |
| | $\mu\text{CCC}_{\text{edge}}^{\text{MSN}}$ | $\mu\text{CCC}_{\text{edge}}^{\text{MIND}}$ |
| **DK** | 0.95 [0.04] | **0.99 [0.01]** |
| **Destrieux** | 0.97 [0.00] | **0.98 [0.00]** |

### Effects of mesh resolution on high-field agreement and ULF reproducibility

**Table S2.**
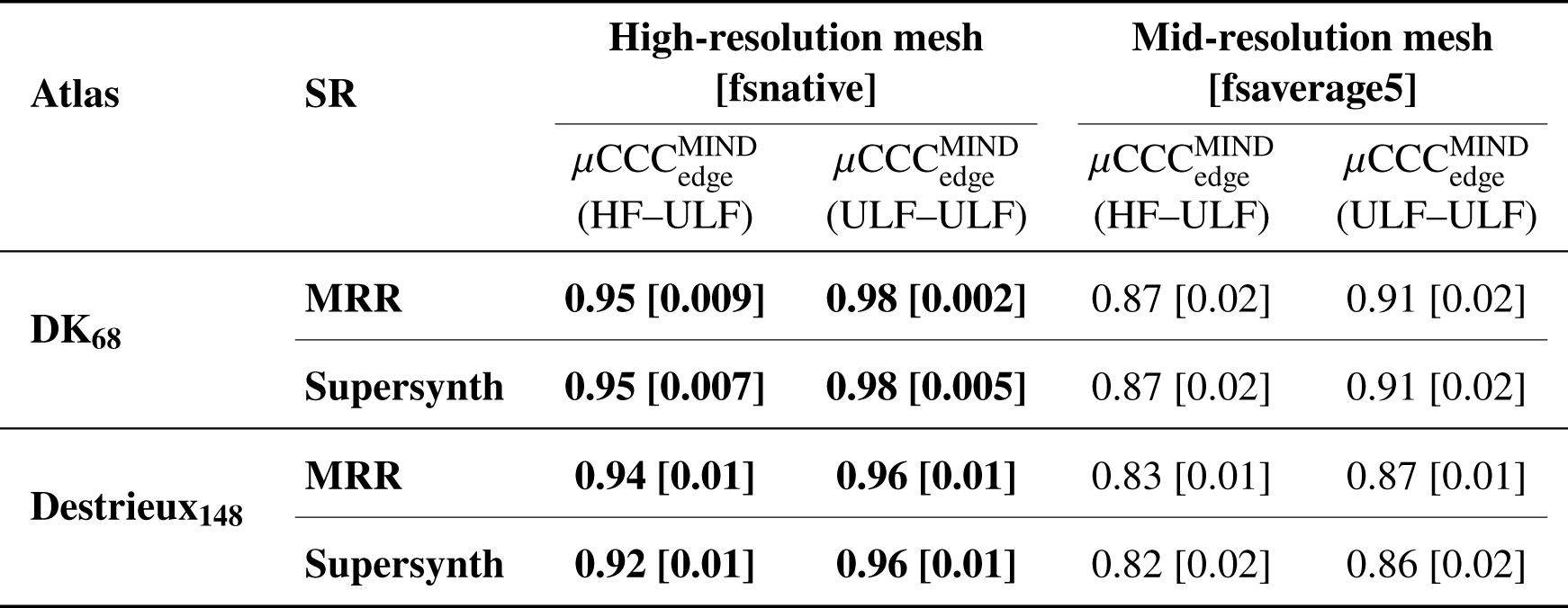
Summary of between-scanner edge-level brain network agreement in MIND networks across mesh resolutions. CCC values are reported as mean [IQR]. Maximum CCC values between mesh resolutions within a category (agreement or reproducibility) are highlighted in bold.

### Agreement and reproducibility captured by ICC

**Figure S1.**
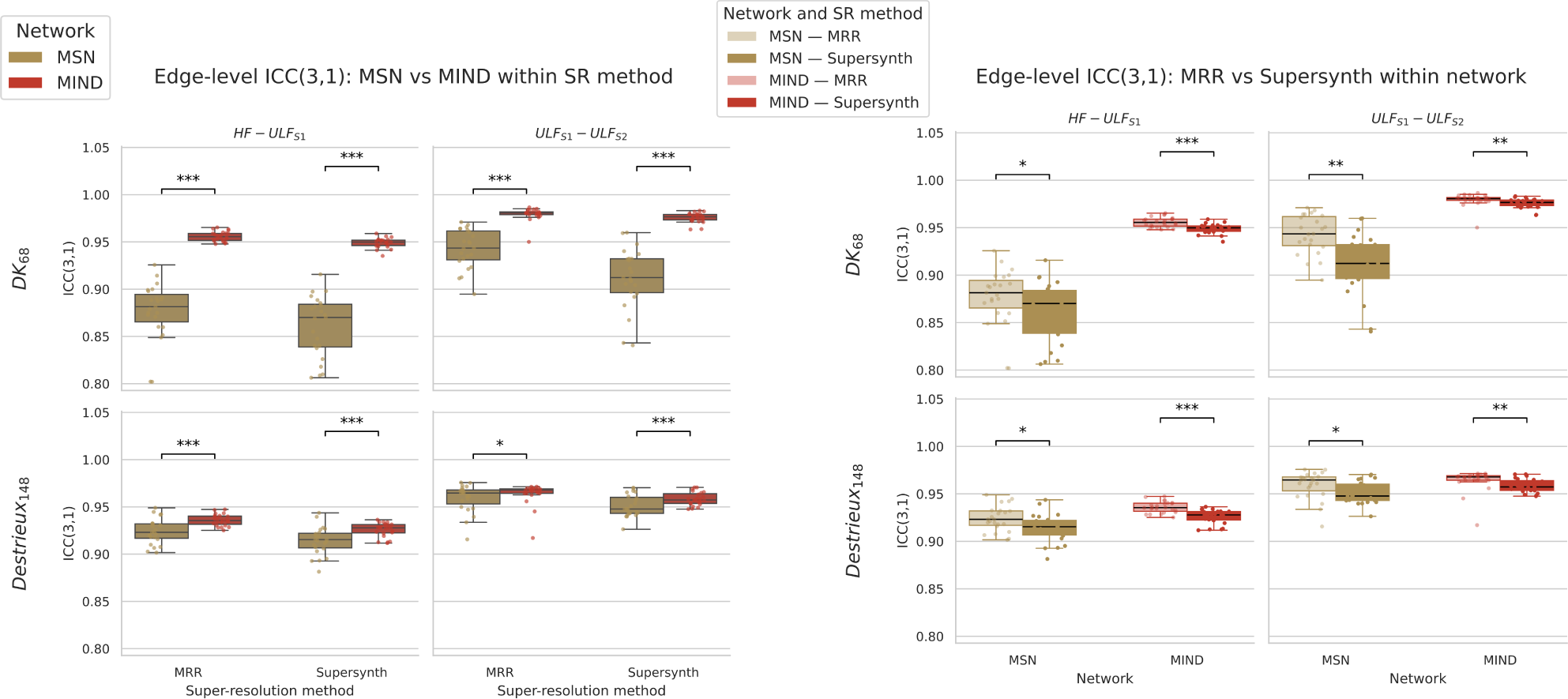
HF-ULF agreement and ULF-ULF reproducibility at the node level for MIND networks across meshes and SR methods, captured by Intraclass Correlation Coefficient (ICC).

### Effects of mesh resolution on identifiability in the Destrieux atlas

**Figure S2.**
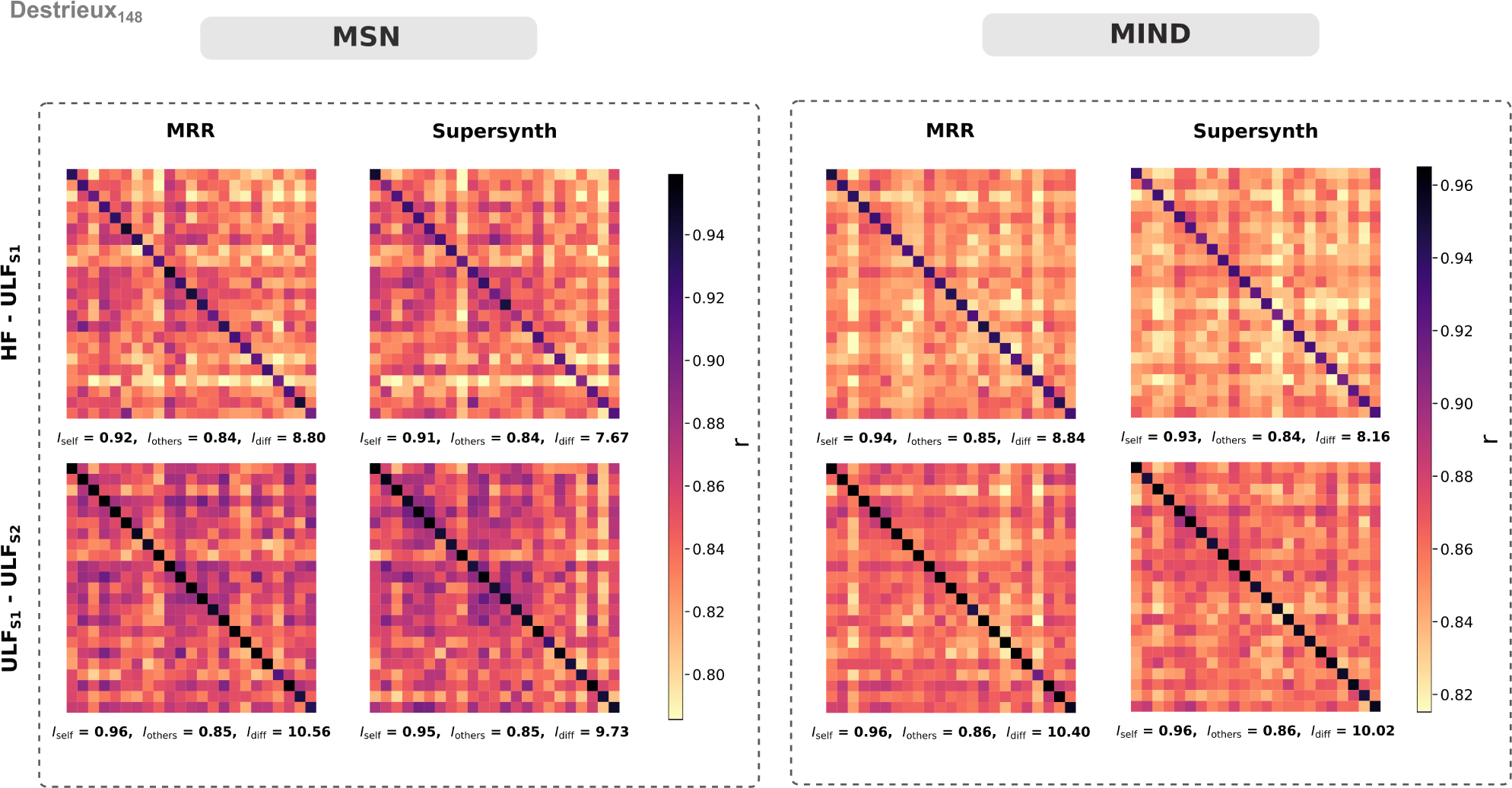
Identifiability plots showing MIND subject-level differentiation across scanners using different mesh resolutions. Results reported using Destrieux_148_ cortical atlas.

## References

Abate, F, A Adu-Amankwah, KA Ae-Ngibise, F Agbokey, VA Agyemang, CT Agyemang, C Akgun, J Ametepe, T Arichi, KP Asante, et al. (2024). “UNITY: A low-field magnetic resonance neuroimaging initiative to characterize neurodevelopment in low and middle-income settings”. In: Developmental Cognitive Neuroscience 69, p. 101397.

Alexander-Bloch, A., J.N. Giedd, and E. Bullmore (2013). “Imaging Structural Co-Variance between Human Brain Regions”. In: Nature reviews neuroscience 14.5, pp. 322–336.

Amico, Enrico and Joaquín Goñi (2018). “The quest for identifiability in human functional connectomes”. In: Scientific reports 8.1, p. 8254.

Baljer, L., N. Bourke, Z. Xie, E. Robinson, and F. Váša (2025). “Multi-input Generalised-Hilbert Mamba for Super-Resolution of Ultra-Low-Field MRI”. In: International Challenge on Ultra-Low-Field MRI Image Enhancement. Cham: Springer Nature Switzerland, pp. 160–172.

Baljer, L., Niall Bourke^1^, Zhenshan Xie, and Emma Robinson (2026). “for Super-Resolution of Ultra-Low-Field”. In: Enhancing Ultra-Low-Field MRI with Paired High-Field MRI Comparisons for Brain Imaging: First International Challenge, ULF-EnC 2025, Held in Conjunction with MICCAI 2025, Daejeon, South Korea, September 23, 2025, Proceedings, p. 160.

Barbas, H. (2015). “General Cortical and Special Prefrontal Connections: Principles from Structure to Function”. In: Annual review of neuroscience 38.1, pp. 269–289.

Barkovich, M.J., Y. Li, R.S. Desikan, A.J. Barkovich, and D. Xu (2019). “Challenges in pediatric neuroimaging”. In: Neuroimage 185, pp. 793–801.

Batalle, D., E. Muñoz-Moreno, F. Figueras, N. Bargallo, E. Eixarch, and E. Gratacos (2013). “Normalization of Similarity-Based Individual Brain Networks from Gray Matter MRI and Its Association with Neurodevelopment in Infants with Intrauterine Growth Restriction”. In: Neuroimage 83, pp. 901–911.

Billot, B., D.N. Greve, O. Puonti, A. Thielscher, K. Leemput, B. Fischl, and J.E. Iglesias (2023a). “SynthSeg: Segmentation of Brain MRI Scans of Any Contrast and Resolution without Retraining”. In: Medical image analysis 86, p. 102789.

Billot, B., C. Magdamo, Y. Cheng, S.E. Arnold, S. Das, and J.E. Iglesias (2023b). “Robust Machine Learning Segmentation for Large-Scale Analysis of Heterogeneous Clinical Brain MRI Datasets”. In: Proceedings of the National Academy of Sciences 120.9, p. 2216399120.

Briski, Ula, Niall J Bourke, Hajer Karoui, Kirsten A Donald, Layla E Bradford, Simone R Williams, Michal R Zieff, Sadia Parkar, Sidra Kaleem, Salman Osmani, et al. (2026). “Deep learning super-resolution of paediatric ultra-low-field MRI without paired high-field scans”. In: Scientific Reports.

Casella, C., A. Leknes, N.J. Bourke, A. Zahra, D. Cromb, D. Barnes, and J. O’Muircheartaigh (2026). MiniMORPH: A Morphometry Pipeline for Low-Field MRI in Infants. Vol. 47. 9. Hoboken, USA: John Wiley & Sons, Inc. 70547 pp.

Cawley, Paul, Francesco Padormo, Daniel Cromb, Jennifer Almalbis, Massimo Marenzana, Rui Teixeira, Sean C Deoni, Emil Ljungberg, Carly Bennallick, Shannon Kolind, et al. (2023). “Development of neonatal-specific sequences for portable ultralow field magnetic resonance brain imaging: a prospective, single-centre, cohort study”. In: EClinicalMedicine 65.

Chen, G., P.A. Taylor, S.P. Haller, K. Kircanski, J. Stoddard, D.S. Pine, and R.W. Cox (2018). “Intraclass Correlation: Improved Modeling Approaches and Applications for Neuroimaging”. In: Human brain mapping 39.3, pp. 1187–1206.

Chetcuti, Karen, C Chilingulo, MS Goyal, L Vidal, NF O’Brien, DG Postels, KB Seydel, and TE Taylor (2022). “Implementation of a low-field portable MRI scanner in a resource-constrained environment: our experience in Malawi”. In: American Journal of Neuroradiology 43.5, pp. 670–674.

Cooper, R., R.A. Hayes, M. Corcoran, K.N. Sheth, T.C. Arnold, J.M. Stein, and M. Jalbrzikowski (2024). “Bridging the Gap: Improving Correspondence between Low-Field and High-Field Magnetic Resonance Images in Young People”. In: Frontiers in neurology 15, p. 1339223.

Davies, A., E. Hutchings, B. Chidiac, M. Garvey, S. Crockford, I. Sebenius, and S. Morgan (2026). MIND the Gap: Methodological Considerations and Guidance for Structural MRI Similarity Network Analysis with MIND.

Deoni, S.C., J. O’Muircheartaigh, E. Ljungberg, M. Huentelman, and S.C. Williams (2022). “Simultaneous High-resolution T2-weighted Imaging and Quantitative T 2 Mapping at Low Magnetic Field Strengths Using a Multiple TE and Multi-orientation Acquisition Approach”. In: Magnetic Resonance in Medicine 88.3, pp. 1273–1281.

Deoni, S.C.L, Muriel MK Bruchhage, Jennifer Beauchemin, Alexandra Volpe, Viren D’Sa, Matthew Huentelman, and Steven CR Williams (2021). “Accessible pediatric neuroimaging using a low field strength MRI scanner”. In: Neuroimage 238, p. 118273.

Dubois, J., M. Alison, S.J. Counsell, L. Hertz-Pannier, P.S. Hüppi, and M.J. Benders (2021). “MRI of the Neonatal Brain: A Review of Methodological Challenges and Neuroscientific Advances”. In: Journal of Magnetic Resonance Imaging 53.5, pp. 1318–1343.

Fan, Q., J. Gao, Y. Wu, Y. Wang, L. Zhang, J. Zhou, and Y. Zhou (2025). Normative Modeling of Brain Morphology Reveals Neuroanatomical Heterogeneity and Biological Subtypes in Major Depressive Disorder. bioRxiv.

Finn, E.S., X. Shen, D. Scheinost, M.D. Rosenberg, J. Huang, M.M. Chun, and R.T. Constable (2015). “Functional Connectome Fingerprinting: Identifying Individuals Using Patterns of Brain Connectivity”. In: Nature neuroscience 18.11, pp. 1664–1671.

Gholam, J., P. Schmid, J. Ametepe, A. Plumley, L. Beltrachini, F. Padormo, and D.K. Jones (2026). “Diffusion Tensor MRI and Spherical-deconvolution-based Tractography on an Ultra-low Field Portable MRI System”. In: Human Brain Mapping 47.2, p. 70454.

Gopinath, K., A. Sorby-Adams, J. Williams-Ramirez, D. Zemlyanker, J. Guo, and D. Hunt (2026). “& Alzheimer’s Disease Neuroimaging Initiative”. In: Human Brain Mapping 47.7, p. 70515.

Goulas, A., H.B. Uylings, and C.C. Hilgetag (2017). “Principles of Ipsilateral and Contralateral Cortico-Cortical Connectivity in the Mouse”. In: Brain Structure and Function 222.3, pp. 1281–1295.

Goulas, A., R. Werner, S.F. Beul, D. Säring, M.V.D. Heuvel, L.C. Triarhou, and C.C. Hilgetag (2016). “Cytoarchitectonic Similarity Is a Wiring Principle of the Human Connectome”. In: BioRxiv 068254.

Han, S., H. Jia, G. Vilaseca, N. Vilaró, F. Duan, Z. Sun, and J. Solé-Casals (2026). “Revealing Structural Brain-Cognition Relationships in Children: A Comparison of Morphometric Similarity and Inverse Divergence Networks”. In: Neuroinformatics 24.1, p. 4.

Hsu, P., E. Marchetto, D.K. Sodickson, P.M. Johnson, and J. Veraart (2025). “Morphological Brain Analysis Using Ultra Low-Field MRI”. In: Human Brain Mapping 46.10, p. 70232.

Iglesias, J.E., B. Billot, Y. Balbastre, C. Magdamo, S.E. Arnold, S. Das, and B. Fischl (2023). “SynthSR: A Public AI Tool to Turn Heterogeneous Clinical Brain Scans into High-Resolution T1-Weighted Images for 3D Morphometry”. In: Science advances 9.5, p. 3607.

Jeffreys, H. (1973). Scientific inference. 3rd. Cambridge University Press.

Jiang, C., Y. He, R.F. Betzel, Y.S. Wang, X.X. Xing, and X.N. Zuo (2023). “Optimizing Network Neuroscience Computation of Individual Differences in Human Spontaneous Brain Activity for Test-Retest Reliability”. In: Network Neuroscience 7.3, pp. 1080–1108.

Kong, Xiang-zhen, Zhaoguo Liu, Lijie Huang, Xu Wang, Zetian Yang, Guangfu Zhou, Zonglei Zhen, and Jia Liu (2015). “Mapping individual brain networks using statistical similarity in regional morphology from MRI”. In: PloS one 10.11, e0141840.

Krupinski, E.A., D. Harris, L.R. Arlinghaus, J. Schlemper, and M. Sofka (2023). “Image Quality Assessment of Advanced Reconstruction Algorithm for Point-of-Care MRI Scanner”. In: Journal of Medical Imaging 10 (Suppl 1), p. 11913.

Li, W., C. Yang, F. Shi, S. Wu, Q. Wang, Y. Nie, and X. Zhang (2017). “Construction of Individual Morphological Brain Networks with Multiple Morphometric Features”. In: Frontiers in neuroanatomy 11, p. 34.

Liang, X., Lianglong Sun, Mingrui Xia, Tengda Zhao, Gaolang Gong, Qiongling Li, Xuhong Liao, Zaixu Cui, Dingna Duan, Chenxuan Pang, et al. (2025). “Dissecting human cortical similarity networks across the lifespan”. In: Neuron 113.19, pp. 3275–3295.

Mangal, Jyoti, Simon Richardson, Yannick Brackenier, Matthew Gardner, Pierluigi Di Cio, Chiara Casella, Shaihan Malik, Jo Hajnal, Martina F Callaghan, Fred Dick, et al. (2026). “Reducing Motion Artifact in High Resolution 7 T MRI Using the Magnetic Resonance Minimal Motion (“MR-MinMo”) Head Stabilization Device”. In: Magnetic Resonance in Medicine.

Montagnese, Marcella, Amir Ebneabbasi, Natalia García-San-Martín, Clara Pecci-Terroba, Rafael Romero-García, Sarah E Morgan, James H Cole, Jakob Seidlitz, Timothy Rittman, and Richard AI Bethlehem (2025). “Structural similarity networks reveal brain vulnerability in dementia”. In: Alzheimer’s & Dementia 21.12, e70973.

Pretzsch, C.M., F. Váša, M. Brammer, L. Brink, M. Potter, W. Mackay, and C. Ecker (2025). “Evaluating Analytic Strategies to Obtain High-Resolution, Vertex-Level Measures of Cortical Neuroanatomy in Children in Low-and Middle-Income Countries”. In: Communications Biology 8.1, p. 918.

Qi, S., S. Meesters, K. Nicolay, B.M. Haar Romeny, and P. Ossenblok (2015). “The Influence of Construction Methodology on Structural Brain Network Measures: A Review”. In: Journal of neuroscience methods 253, pp. 170–182.

Raznahan, A., Jason P Lerch, Nancy Lee, Dede Greenstein, Gregory L Wallace, Michael Stockman, Liv Clasen, Phillip W Shaw, and Jay N Giedd (2011). “Patterns of coordinated anatomical change in human cortical development: a longitudinal neuroimaging study of maturational coupling”. In: Neuron 72.5, pp. 873–884.

Ringshaw, JE, Michal R Zieff, Niall J Bourke, Chiara Casella, Layla E Bradford, Simone R Williams, Donna Herr, Marlie Miles, Carly Bennallick, Khula South Africa Data Collection Team Davel Lauren Pan Tracy Mhlakwaphalwa Tembeka Methola Bokang Nkubungu Khanyisa Knipe Candice Madi Zamazimba Mlandu Nwabisa Gulwa Ringie Madikane Pamela, et al. (2026). “Antenatal maternal anaemia and infant brain structure: high-field (3 T) and ultra-low-field (64 mT) MRI findings from South Africa”. In: Brain Communications 8.5, fcag318.

Romero-Garcia, R., M. Atienza, L.H. Clemmensen, and J.L. Cantero (2012). “Effects of Network Resolution on Topological Properties of Human Neocortex”. In: Neuroimage 59.4, pp. 3522–3532.

Romero-Garcia, R., Kirstie J Whitaker, František Váša, Jakob Seidlitz, Maxwell Shinn, Peter Fonagy, Raymond J Dolan, Peter B Jones, Ian M Goodyer, Edward T Bullmore, et al. (2018). “Structural covariance networks are coupled to expression of genes enriched in supragranular layers of the human cortex”. In: Neuroimage 171, pp. 256–267.

Sebenius, I., Lena Dorfschmidt, Jakob Seidlitz, Aaron Alexander-Bloch, Sarah E Morgan, and Edward Bullmore (2025). “Structural MRI of brain similarity networks”. In: Nature Reviews Neuroscience 26.1, pp. 42–59.

Sebenius, I., Jakob Seidlitz, Varun Warrier, Richard AI Bethlehem, Aaron Alexander-Bloch, Travis T Mallard, Rafael Romero Garcia, Edward T Bullmore, and Sarah E Morgan (2023). “Robust estimation of cortical similarity networks from brain MRI”. In: Nature neuroscience 26.8, pp. 1461–1471.

Seidlitz, J., František Váša, Maxwell Shinn, Rafael Romero-Garcia, Kirstie J Whitaker, Petra E Vértes, Konrad Wagstyl, Paul Kirkpatrick Reardon, Liv Clasen, Siyuan Liu, et al. (2018). “Morphometric similarity networks detect microscale cortical organization and predict inter-individual cognitive variation”. In: Neuron 97.1, pp. 231–247.

Sorby-Adams, A.J., J. Guo, P. Laso, J.E. Kirsch, J. Zabinska, A.L. Garcia Guarniz, and W.T. Kimberly (2024). “Portable, Low-Field Magnetic Resonance Imaging for Evaluation of Alzheimer’s Disease”. In: Nature Communications 15.1, p. 10488.

Stockbridge, M.D., R. Wang, V. Neal, I. Diaz-Carr, A.E. Hillis, and A.V. Faria (2026). “Reproducibility of Volumetric Analysis Using Ultra-Low-Field Portable Magnetic Resonance Imaging”. In: Aperture Neuro 6.

Tapp, A., C. Zhao, H.R. Roth, J. Tanedo, S.M. Anwar, N.J. Bourke, and M.G. Linguraru (Oct. 2024). “Super-Field MRI Synthesis for Infant Brains Enhanced by Dual Channel Latent Diffusion”. In: International Conference on Medical Image Computing and Computer-Assisted Intervention. Cham: Springer Nature Switzerland, pp. 444–454.

Tranfa, M., M. Petracca, M. Moccia, A. Scaravilli, F. Barkhof, V. Brescia Morra, and G. Pontillo (2025). “Conventional MRI-Based Structural Disconnection and Morphometric Similarity Networks and Their Clinical Correlates in Multiple Sclerosis”. In: Neurology 104.4, p. 213349.

Váša, F., C. Bennallick, N.J. Bourke, F. Padormo, L. Baljer, U. Briski, and S.C. Williams (2025). “Ultra-Low-Field Brain MRI Morphometry: Test–Retest Reliability and Correspondence to High-Field MRI”. In: Imaging Neuroscience 3, pp. –.

Váša, F., J. Seidlitz, Rafael Romero-Garcia, Kirstie J Whitaker, Gideon Rosenthal, Petra E Vértes, Maxwell Shinn, Aaron Alexander-Bloch, Peter Fonagy, Raymond J Dolan, et al. (2018). “Adolescent tuning of association cortex in human structural brain networks”. In: Cerebral Cortex 28.1, pp. 281–294.

Vokhiwa, M., K. Chetcuti, F. Lange, N.J. Bourke, L. Randall, S. Greenstein, and E. Umar (2026). “Feasibility, Image Quality, and Analytic Usability of Mobile 64 mT Ultra-Low-Field MRI for Infant Brain Imaging at 3 and 12 Months in Southern Malawi”. In: Journal of Magnetic Resonance Open 100225.

Wang, Y., D. Zhu, L. Zhao, X. Wang, Z. Zhang, B. Hu, and W. Zheng (2024). “Profiling Cortical Morphometric Similarity in Perinatal Brains: Insights from Development, Sex Difference, and Inter-Individual Variation”. In: NeuroImage 295, p. 120660.

Yao, G., J. Luo, T. Zou, J. Li, S. Hu, L. Yang, and P. Liu (2024). “Transcriptional Patterns of the Cortical Morphometric Inverse Divergence in First-Episode, Treatment-Naïve Early-Onset Schizophrenia”. In: Neuroimage 285, p. 120493.

Yao, Z., Yuanchao Zhang, Lei Lin, Yuan Zhou, Cunlu Xu, Tianzi Jiang, and Alzheimer’s Disease Neuroimaging Initiative (2010). “Abnormal cortical networks in mild cognitive impairment and Alzheimer’s disease”. In: PLoS computational biology 6.11, e1001006.

Zalesky, A., A. Fornito, I.H. Harding, L. Cocchi, M. Yücel, C. Pantelis, and E.T. Bullmore (2010). “Whole-Brain Anatomical Networks: Does the Choice of Nodes Matter?” In: Neuroimage 50.3, pp. 970–983.

Zimmermann, J., J. Griffiths, M. Schirner, P. Ritter, and A.R. McIntosh (2018). “Subject Specificity of the Correlation between Large-Scale Structural and Functional Connectivity”. In: Network Neuroscience 3.1, pp. 90–106.

